# Fine-scale genetic structure and population-specific clinically relevant variants in the indigenous tribal populations of India

**DOI:** 10.64898/2026.01.11.26343796

**Authors:** Sudeshna Datta, Deepak Jena, Arup Ghosh, Viplov Kumar Biswas, Sudarshana Jena, Mamuni Swain, Vinay More, Soumendu Mahapatra, Rasmita Mishra, Bhawna Gupta, ILS Flagship Consortium, Analabha Basu, Punit Prasad, Sunil Kumar Raghav

**Author notes:** First author (Sudeshna Datta). Corresponding author: Contact email ID for the Corresponding author.

## Abstract

India harbors one of the most complex demographic and genetic landscapes globally, comprising more than 4600 endogamous ethnolinguistic groups shaped by multiple waves of migration, prolonged isolation and deep social stratification. Odisha, an eastern state of India, is home to 62 indigenous tribal groups, including 13 Particularly Vulnerable Tribal Groups (PVTGs), many of which remain genetically understudied. These populations provide a unique opportunity to explore fine-scale population structure and population-specific variomes in underrepresented South Asian populations. We generated high-coverage whole-exome sequencing (WES) of 13 tribal groups (n = 515) from Odisha, collectively termed as OdiTribe. We identified 625,596 single-nucleotide variants (SNVs), of which 42.51% were rare and 5.54% (n=34,667) were novel. Population genetic analyses revealed clear substructure within the OdiTribe, along with pronounced population-specific drift in the Juang tribe. The Juang and Bonda tribes exhibited significant shared genetic drift with the Nicobarese, as indicated by outgroup F3 statistics, suggesting ancestral connections. Approximately 43% of the individuals had high inbreeding coefficients (>= 0.0156), and 14.2% carried ROH segments > 8 Mb, indicating recent consanguinity. Functional annotation further uncovered population-specific and clinically relevant pathogenic variants in *HBB*, *HBD, BTD* and *BRCA2* genes, as well as multiple high-confidence homozygous loss-of-function (HC-LoF) variants across the dataset. Our study provides exome-wide insights into genetic diversity, endogamy and population-specific pathogenic variants among endogamous tribal populations. These findings expand the catalogue of South Asian genomic variation and also highlight the importance of including isolated indigenous groups in global population and medical genomics research.

## Introduction

India, home to approximately 1.44 billion people, is a country of remarkable diversity, comprising over 4,600 distinct endogamous ethno-linguistic groups ^1^. These groups can be classified into four major linguistic families, i.e., Indo-European (IE), Dravidian (DR), Austroasiatic (AA) and Tibeto-Burman (TB), each associated with unique demographic histories and geographic distributions. Genome-wide studies on autosomal markers have revealed that present-day Indians are a result of admixture from multiple ancestral components: the Ancestral North Indian (ANI), Ancestral South Indian (ASI), Ancestral Austroasiatic (AAA) and Ancestral Tibeto-Burman (ATB) ^2^. Socially, Indian populations are broadly stratified into castes and tribes, with varying proportions of ANI, ASI, AAA and ATB ancestry. The populations, irrespective of caste or tribal background, are also known to practice consanguineous marriages. Thus, a combination of long-term endogamy and consanguinity has resulted in an increased occurrence of recessive disease risk among the populations ^3^,^4^. Together, India’s complex demographic history and unique social stratification make it an invaluable cohort for understanding population-specific genetic variation, demographic dynamics and disease risk architecture.

Approximately 705 groups are officially recognized as the “Scheduled Tribes”, comprising 8.6% of the total population (Government of India, Census 2011). The origin of these tribes has long been debated among anthropologists, linguists and geneticists but over the past decade, research integrating uniparental markers, genome-wide SNP arrays and whole-genome sequencing (WGS) has advanced our understanding of the origin and migration of humans in South Asia as well as the genetic diversity of tribal and caste populations ^2,4–9^. A Y-chromosome haplogroup study on 12 Dravidian tribal and 19 caste endogamous populations revealed high proportion of Indian autochthonous Y-chromosomal haplogroups (H-M69, F-M89, R1a1-M17, L1-M27, R2-M124, and C5-M356) comprising of 81% of all male lineages and suggesting a low genetic influence from west Eurasian migration ^5^. Studies using mitochondrial DNA (mtDNA) revealed that most of the maternal lineages in India belong to the M macrohaplogroup followed by U and R ^6,7^. Although population genetic studies have been conducted across several Indian states, Odisha remains comparatively understudied, with a few being an autosomal microsatellite and uniparental marker investigations available for a few caste and tribe groups ^10–12^. A comprehensive genomic characterization is essential to better understand the population history and genetic landscape of Odisha’s population.

Odisha, an eastern state of India, has the third-largest tribal population and is home to 62 tribal groups, constituting 22.8% of its population ^13^. The state is also home to 13 of India’s 75 Particularly Vulnerable Tribal Groups (PVTGs), which are among the most marginalized communities in the nation ^13,14^. PVTGs are identified based on four features: they live in isolation and are largely unaffected by the developmental process, have a lower literacy rate, a pre-agricultural level of technology and a stagnant population ^14^. Linguistically, Odisha’s tribes belong to three families, i.e. IE, AA, and DR. Among them, Kharia, Juang, Gadaba, Ho, and Saora are among the few most ancient tribes belonging to the AA linguistic family, whereas Paraja, Oraon, and Kondh belong to the DR linguistic group ^10^. Genetic studies using uniparental and autosomal markers have shown that Odisha’s tribal groups exhibit deep-rooted genetic continuity and long-term isolation. In contrast, caste groups display genetic affinity with North Indian caste groups ^10,12^.

The tribal populations across India experience a higher disease burden compared to the other caste groups, largely due to geographical isolation and socio-economic marginalization, which creates systemic barriers to accessing healthcare systems ^13^. In Odisha, health outcomes among tribal communities are significantly poorer than national averages, with high rates of malnutrition, poor sanitation and inadequate access to maternal and child healthcare services ^13,15^. Epidemiological studies reveal a high prevalence of malnutrition, infectious diseases such as malaria, tuberculosis, and genetic disorders like G6PD deficiency, sickle cell anemia, and thalassemia (ICMR Bulletin, October 2003) ^16^. The overall prevalence of sickle cell anemia among PVTGs is around 3.3%, with variations among tribal groups ^16^. The elevated burden of hereditary disorders aligns with the history of strong endogamy and restricted gene flow observed in prior genetic studies in other populations ^2^,^17^.

In this study, we performed whole-exome sequencing (WES) of 634 healthy individuals from 13 tribal groups in Odisha to understand their genetic diversity and differentiation. By integrating with the GenomeAsia100K dataset ^18^, we explored the population structure, admixture patterns, inbreeding and functional variant profile. This represents one of the largest WES datasets generated from Indian tribal populations to date. Our study provides new insights into the genetic differentiation, ancestral composition, and population-specific variants within Odisha’s tribal population, thus shedding light on their demographic history and potential genetic predisposition to diseases among some of the underrepresented populations of India.

## Materials and Methods

### Sample collection

The study samples comprised 634 self-declared healthy participants collected from six districts of Odisha: Sundergarh, Keonjhar, Kandhamal, Nabarangpur, Malkangiri, and Kandhamal. All the participants provided written consent and responded to questions about socio-demographic, lifestyle and health-related factors using a detailed questionnaire. Additionally, anthropometric measurements were also obtained. Approximately 10 ml of blood was collected from each individual, which was processed for biochemical tests and DNA isolation. All the sample and data collection methods were conducted as per ethical guidelines and regulations after receiving due approval (IEC no: 87/HEC/19) from the Institutional Ethics Committee (IEC) at the Institute of Life Sciences.

### DNA extraction and library preparation

DNA was extracted from the blood samples using the MasterPure Blood Purification kit (Lucigen), and quality was checked using Nanodrop, followed by accurate quantification with a Qubit 4.0 fluorometer. Whole-exome sequencing libraries were prepared using the Agilent V6+UTR protocol and sequenced to get a coverage of 100X in four batches on a Novaseq 6000 machine with 150bp paired-end reads.

### Variant calling pipeline

The base call files were demultiplexed using the bcl2fastq software, and the quality of the fastq files was checked using the FASTQC tool (https://www.bioinformatics.babraham.ac.uk/projects/fastqc/). To reduce false positives or error rates in the raw reads, the adapter and low-quality bases were removed using fastp (version 0.39), a quality control software tool for sequencing data. All high-quality paired-end reads were mapped against the hg38 human reference genome using the BWA-MEM aligner (version 0.7.17) and samples less than 10X coverage were not included in downstream analysis. The Picard MarkDuplicates tool (version 2.26.5) (http://broadinstitute.github.io/picard) was used to mark the PCR duplicate reads, which were subsequently excluded from downstream analysis. Base quality score recalibration and variant calling were performed using GATK (version 4.1.2.0) (Genome Analysis ToolKit) according to GATK Best Practices recommendations ^19^. In brief, HaplotypeCaller was used to call variants for each sample, which were then aggregated using GenomicsDBImport in a chromosome-wise manner. Following this, we performed joint genotyping using GenotypeGVCFs. For the selection of high-quality variants, we performed VQSR. Variant recalibration was applied by the ApplyRecalibration of GATK using a tranche sensitivity of 99% for both SNVs and indels. Further quality control was restricted to only SNVs. We have filtered out SNVs using PLINK v1.9 (http://pngu.mgh.harvard.edu/purcell/plink/) ^20^ with missingness rates higher than 1% and deviation from Hardy-Weinberg equilibrium with a p-value of < 0.00000001. Mean target coverage and variant summary statistics of the samples were evaluated using CollectHSmetrics and BCFtools stats ^21^, respectively (**Supplementary Figure 1A and 1B**).

### Identification of first-degree relatives

Relatedness analysis was performed using KING ^22^. PLINK v1.9 ^20^ was used to convert the VCF file to the required input format. First-degree relatives were identified and excluded using the kinship coefficient threshold of 0.177 - 0.354 as described in the manual. As a result, 99 related individuals were removed, leaving 515 unrelated samples for downstream analyses.

### Population Structure Analysis

To understand the genetic structure of the Odisha tribe (OdiTribe) population, we generated three datasets. First, we merged the OdiTribe populations with those from the GenomeAsia 100K cohort ^18^: West Eurasia (n = 111), Southeast Asia (n = 333) and South Asian (n = 604) populations. For the GenomeAsia dataset, we lifted over the hg19 variants to hg38 using the hg19 to hg38 chain file downloaded from the UCSC website. Following this, we extracted the target regions corresponding to the probe sets used in the exome sequencing and combined them with the OdiTribe (global dataset; n=1,563). Second, we generated the regional dataset by merging populations with the closest relationship to the OdiTribe populations, as determined by Wright’s fixation index (FST) **(Supplementary Figure 3C)**. This dataset comprises Indian tribal populations from the GenomeAsia 100K cohort and the OdiTribe populations (n = 702). Lastly, to understand the substructure within the OdiTribe populations, we used the filtered OdiTribe dataset (n = 515). For all the VCF dataset files, they were converted to PLINK binary format using PLINK v1.9 (ref). The binary files were then merged, and variants were filtered for minor allele frequency (MAF <0.05) and pruned for linkage disequilibrium (LD); sliding window = 50 kb, step size = 5 and r2 threshold = 0.5. For visualisation, we used various R (v4.1.0) packages: ggplot2 (v2.3.5), reshape (v1.4.4), dplyr (v1.1.2) and stringr (v1.5).

### Principal Component Analysis

We used the EIGENSOFT SmartPCA ^23^, first to explore variation in a global context by using the global dataset, second, to understand variation in a regional context using the regional dataset and third, to understand the variation within OdiTribe populations.

### Admixture analysis

To assess the ancestral genetic composition in the OdiTribe populations, we performed model-based clustering using ADMIXTURE v1.3.0 ^24^. Analysis with k from 2 to 18 was performed for the global dataset. For the OdiTribe dataset, admixture analysis was performed for k values ranging from 2 to 8. The lowest cross-validation (CV) error parameter was obtained at k = 18 and k = 6 for the global and OdiTribe datasets, respectively **(Supplementary Figure 3A and Supplementary Figure 6A)**.

### Wright’s fixation index

The degree of differentiation among the populations included in the global and regional datasets was evaluated with Fst values produced by Weir and Cockerham estimation which is included in the EIGENSOFT SmartPCA ^23^.

### Maximum likelihood tree construction

We evaluated the genetic drift for the regional dataset by constructing a maximum likelihood (ML) tree using TreeMix v.1.3 software ^25^.

### F3 outgroup analysis

The Admixr package was used to calculate outgroup F3 statistics ^26^. To infer gene flow in OdiTribe populations, F3 statistics were used as F3 (Yoruba; OdiTribe, X), where X is the Indian tribal population groups, and OdiTribe groups represent the 13 tribal groups.

### Inbreeding coefficient and runs of homozygosity (ROH)

Plink --het ^20^ was used to determine the inbreeding coefficients (Fplink). We detected several individuals with negative Fplink values, which could reflect a recent admixture of previously diverse populations or biased variant sampling ^27^. The ROH analysis was performed using the plink --homozyg option with optimised parameters for WES ^17^,^28^. We used all the default parameters while setting the sliding window to 50kb without any heterozygous sites (--homozyg-snp 50, --homozyg-window-het 0). Only ROH segments with a length of 1 Mb were selected. Following the classification method used in a recent paper ^17^ we too binned the ROHs into five classes: Class A (1 Mb – 2 Mb), Class B (2 Mb – 4 Mb), Class C (4 Mb – 8 Mb), Class D (8 Mb –16 Mb) and Class E (>16 Mb).

### Functional and Disease annotation

Variants were annotated using various tools such as Annovar ^29^, SnpEff ^30^ and VEP ^31^. The high-confidence (HC) and low-confidence (LC) predicted loss-of-function (LoF) variants (stop codon, splice site, frameshift mutation) were detected using the LOFTEE ^32^ plugin in VEP ^31^. Variants are also annotated by the Annovar tool ^29^ using data from dbnsfp35a ^33^, which includes SIFT ^34^, Polyphen2 ^35^, Mutation Taster ^36^, CADD ^37^, GERP++ ^38^ and population data from gnomAD v.4.0 ^39^ and 1000 Genome phase3 data ^40^. We also annotated our variants against the ClinaVar version (20240917) ^41^ to identify pathogenic/Likely Pathogenic (P/LP) in the same run using a VCF file. We also compared the allele frequency of OdiTribe variants against GenomeAsia ^18^, gnomAD ^39^ and 1000 Genome ^40^ cohort for identifying population-specific disease-causing variants present at high frequencies. Both heterozygous and homozygous variants were reported. Over-representation analysis was performed using the ConsensusPathDB database ^42^ to identify overrepresented biological pathways. Additionally, the variants were classified as “Novel” if they had no record in the dbSNP build 156 ^43^ and gnomAD v4.0 ^39^ databases and identified population-specific patterns. Further, we assessed the functional impact of novel missense variants using two predictive scoring systems: REVEL (Rare Exome Variant Ensemble Learner) ^44^ and CADD (Combined Annotation Dependent Depletion) ^37^. REVEL combines scores from 13 different computational tools (MutPred, FATHMM, VEST, SIFT, PolyPhen-2, MutationTaster and others) to assess the deleterious effect of the variant. Missense variants with REVEL scores >= 0.644 and CADD scores >= 20 were considered as deleterious.

The American College of Medical Genetics (ACMG) SF v3.2 ^45^ recommended actionable 81 genes were screened. Fisher’s exact test was used to calculate the variations that were significantly high in frequency in the OdiTribe samples compared to the global datasets.

Further, we annotated all variants that were identified in the OdiTribe dataset against ClinVar v.20240917 and Online Mendelian Inheritance in Man (OMIM) ^46^. Only disease-causing pathogenic and likely-pathogenic mutations in ClinVar were used for further analysis. Inheritance types of phenotypes were extracted from the OMIM database. Additionally, only pathogenic and likely pathogenic variants in ACMG-recommended actionable genes were reported.

## Results

### Whole-exome sequencing reveals the genetic variation landscape of the OdiTribe

We generated high-coverage (median depth of coverage 87x) **(Supplementary Figure 1A)** whole-exome sequencing (WES) data from 634 indigenous individuals from Odisha, collectively referred to as OdiTribe here in the manuscript. These individuals belong to 13 indigenous tribal groups, including 4 Particularly Vulnerable Tribal Groups (PVTGs) and spanning 3 major linguistic families **(Figure 1A)**. Among them, 46.7% spoke Austroasiatic (AA; n = 296), 33.9% Dravidian (DR; n = 215), and 19.4% Indo-European (IE; n = 123) languages (**Supplementary Figure 1B**). Approximately 60% of the participants were females and 40% males, yielding a female-to-male ratio of about 3:2, with median ages 40 and 41 years for females and males, respectively **(Supplementary Figure 1C)**. After filtering the samples based on the sequencing data quality and removal of first-degree related individuals (see methods for details), 515 exome samples were left for downstream analysis **(Supplementary Figure 1D-E, Table 1)**. Following the GATK best practice workflow ^19^, a total of 6,84,841 high-confidence bi-allelic variants consisting of 6,25,596 single-nucleotide variants (SNVs) (91.3%) and 59,245 indels (8.7%) were identified. Only SNVs were considered for downstream analyses; therefore, the terms “SNV” and “variant” are used interchangeably throughout the manuscript. A large proportion, 33.63% of the variants, were singletons (Allele Count, AC = 1). Of the total SNVs, 16.15% were unreported in dbSNP (v156) ^43^ and gnomAD (v4.0) ^39^ databases. The unreported SNVs included 5.54% non-singletons and 10.61% singletons **(Figure 1B)**. For functional analyses, we defined these unreported non-singleton SNVs as novel variants. The presence of these notable fractions of novel variants in the OdiTribe dataset underscores the existence of population-specific variation that remains underrepresented in global reference datasets. Further, based on the minor allele frequency (MAF) distribution, the identified SNVs were categorized as rare (MAF < 0.01; n = 2,31,625), uncommon (0.01 < MAF < 0.05; n = 65,214) and common (MAF >0.05; n = 1,18,344) SNVs **(Figure 1C)**.

**Figure 1.**
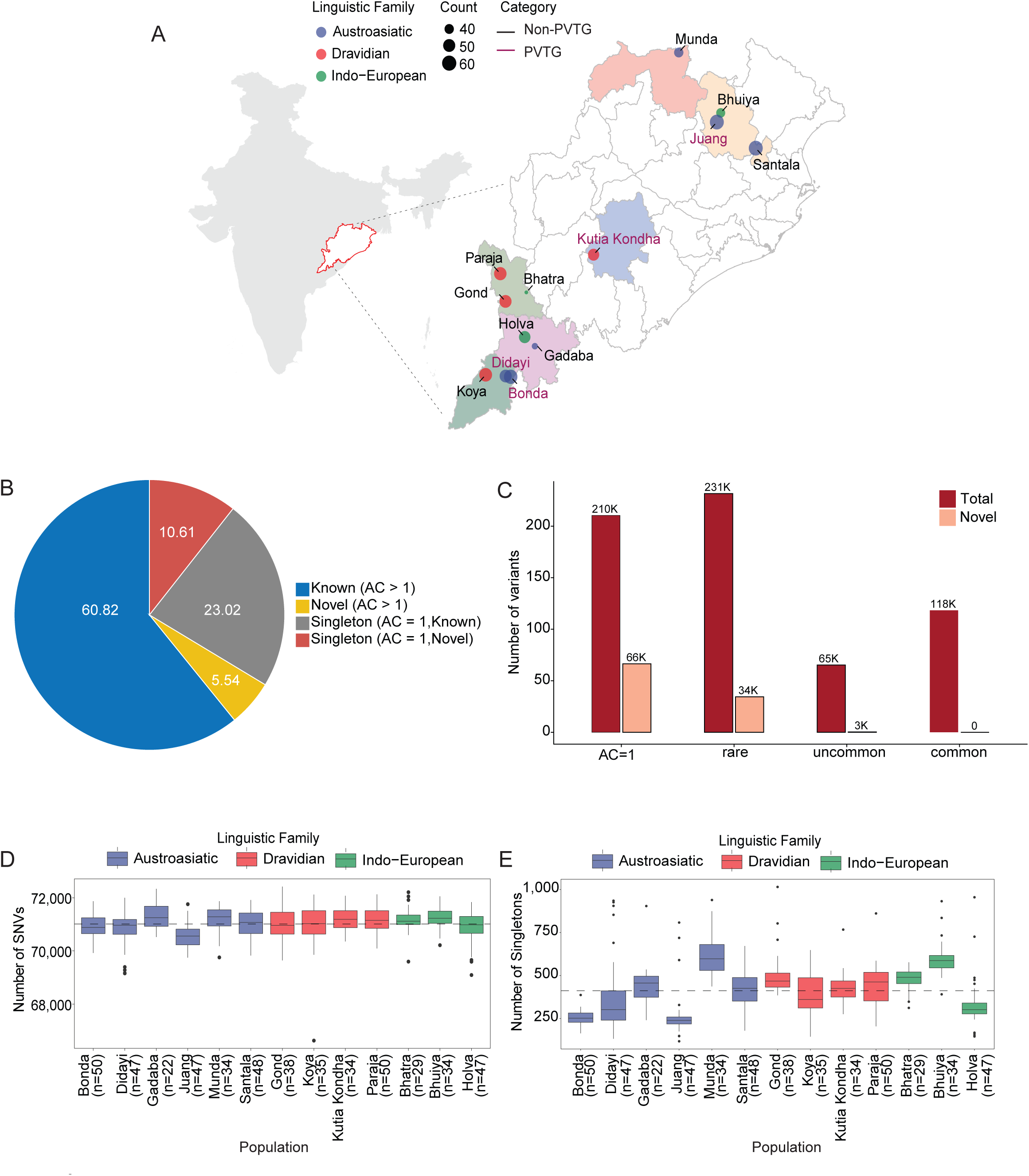
Sampling overview and summary of high-quality variants. A. Map of Odisha showing the sampling sites. Each point denotes a tribal group, colored by linguistic family, and the size indicates the sample count. B. Percentage of novel and known SNVs classified by singletons (AC = 1) and non-singletons (AC > 1). C. Distribution of total and novel SNVs across MAF categories: rare (MAF < 0.01), uncommon (0.01 > MAF < 0.05) and common (MAF > 0.05). AC = 1 denotes singletons. D. Distribution of total SNVs across the OdiTribe groups. Groups are colored according to their linguistic family. E. Distribution of singletons across the OdiTribe groups. Groups are colored according to their linguistic family.

**Table 1:**
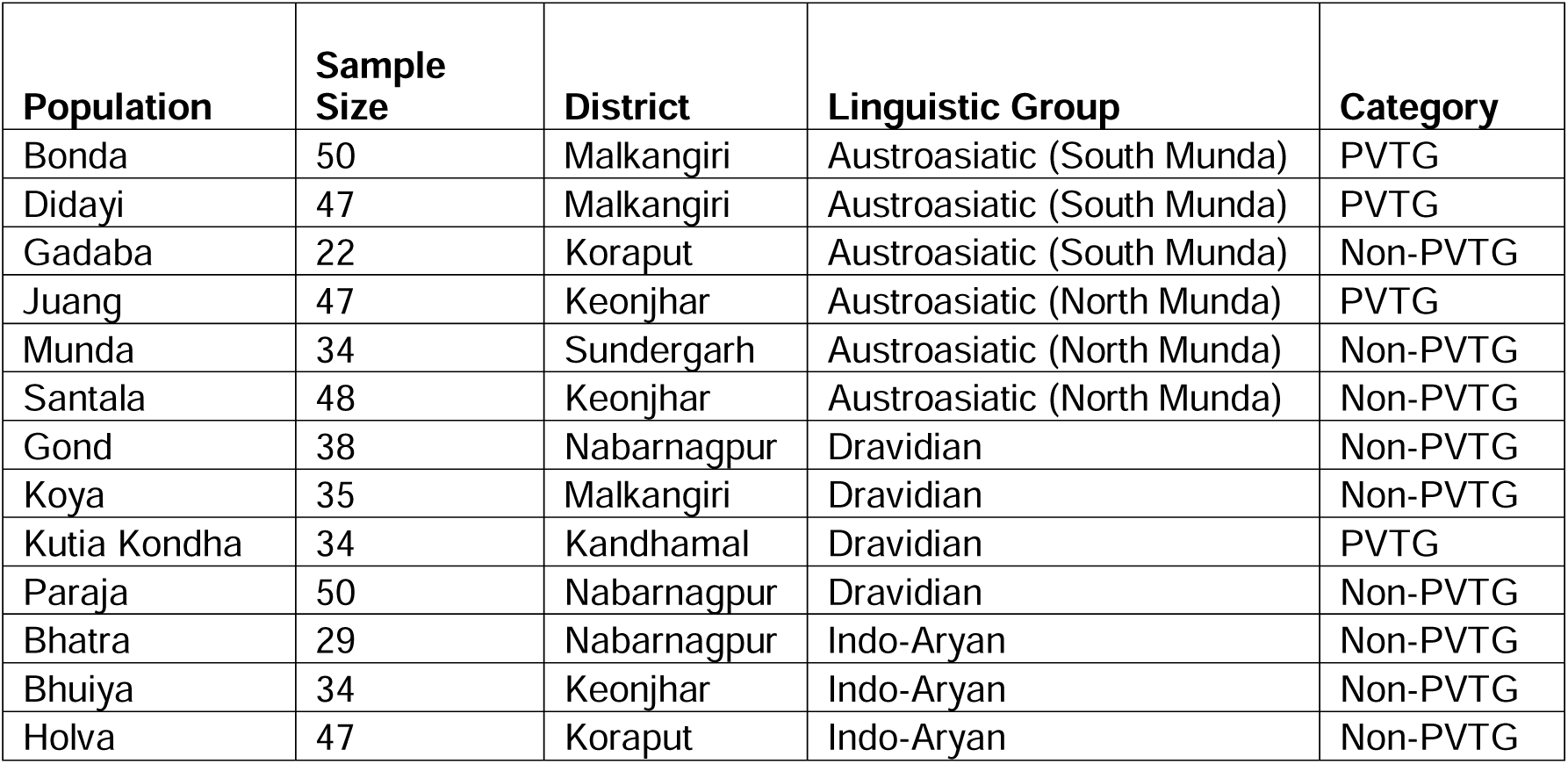
Sample details of the OdiTribe.

The average number of SNVs captured per individual was around 70,000, with a median of 71,055 (IQR: 70679 - 71393). Individuals from the Gadaba tribe exhibited the highest number of SNVs (mean = 71302.05, sd = 462.7), and the Juang tribe exhibited the lowest (mean = 70577.57, sd = 451.39). Other groups, such as Bhuiya and Munda, showed a relatively higher number of SNVs (mean = 71234.65, sd = 389.7 and 71188.29, sd =478.27, respectively) while Didayi and Holva exhibited a lower SNV burden (mean = 70833.19 ± 660.82 and 70908.81 ± 568.58, respectively) **(Figure 1D, Supplementary Table 1).** Similar to the distribution of total SNVs, Munda, Bhuiya, and Gadaba showed higher singleton counts, while Juang, Bonda, and Holva carried comparatively fewer counts **(Figure 1D, Supplementary Table 1)**. AA speaking groups harbored a smaller number of SNVs than DR and IE groups, whereas DR and IE groups did not differ much, and the same was observed in singleton distribution **(Supplementary Figure 1F, Supplementary Table 2)**. Among the common variants, approximately 94% were shared across all three linguistic groups, while ∼5% were rare in at least one. In contrast, ∼3% of the rare variants were common in at least one linguistic group **(Supplementary Table 3)**.

### Population structure and genetic affinities of the OdiTribe

We combined the OdiTribe dataset with West Eurasian, South Asian, and Southeast Asian populations (global dataset) from the GenomeAsia 100K dataset ^18^ to assess the genetic differentiation of the OdiTribe groups in a global context **(Figure 2A; Supplementary Figure 2A)**. Principal component analysis revealed the distinction of Southeast Asia from West Eurasia, and South Asian populations across PC1 and PC2 distinguished West Eurasian from the rest **(Figure 2A)**. To understand the genetic diversity of the OdiTribe with respect to other mainland Indian populations, we classified the Indian samples from the GenomeAsia100K dataset ^18^ into seven ethnolinguistic and social categories: Indo-European Caste (IE-C), Indo-European Tribe (IE-T), Dravidian Caste (DR-C), Dravidian Tribe (DR-T), Austroasiatic Tribe (AA-T) and Tibeto-Burman Tribe (TB-T). Since Andamanese and Nicobarese exhibit distinct ancestry compared to mainland Indians, they were not included in these categories. In concordance with previous studies ^3^,^4^, the Indian groups were spread within the South Asian cline extending from IE-C groups towards DR-T groups along the PC2 axis **(Figure 2A)**. The TB-T groups overlapped with the Southeast Asian groups sharing the East Asian ancestry ^47^. The OdiTribe individuals clustered tightly at the distal end of this cline with a slight shift along the first component, grouping with other AA and DR tribes. We further grouped by the geographic origin of the Indian samples – North, South, East, Central and Northeast India. As expected, the IE-C groups were positioned toward the northern end of the cline, and the DR groups (both caste and tribe) were along the southern end of the cline, overlapping with western Indian groups **(Supplementary Figure 2B)**. The OdiTribe, representing eastern Indian tribal populations, occupied the southern end of the cline and clustered closely with DR-T and AA-T groups from central, eastern and southern India **(Supplementary Figure 2B)**. Pairwise Fst estimates further supported these observations, showing that the OdiTribe groups are genetically closest to the DR-T, AA-T and IE-T groups and more differentiated from the caste populations **(Figure 2B).** Further, when examined at the individual population level, fine structure was observed thus highlighting the intra-population genetic affinities and differentiation among OdiTribe groups **(Supplementary Figure 2C)**.

**Figure 2.**
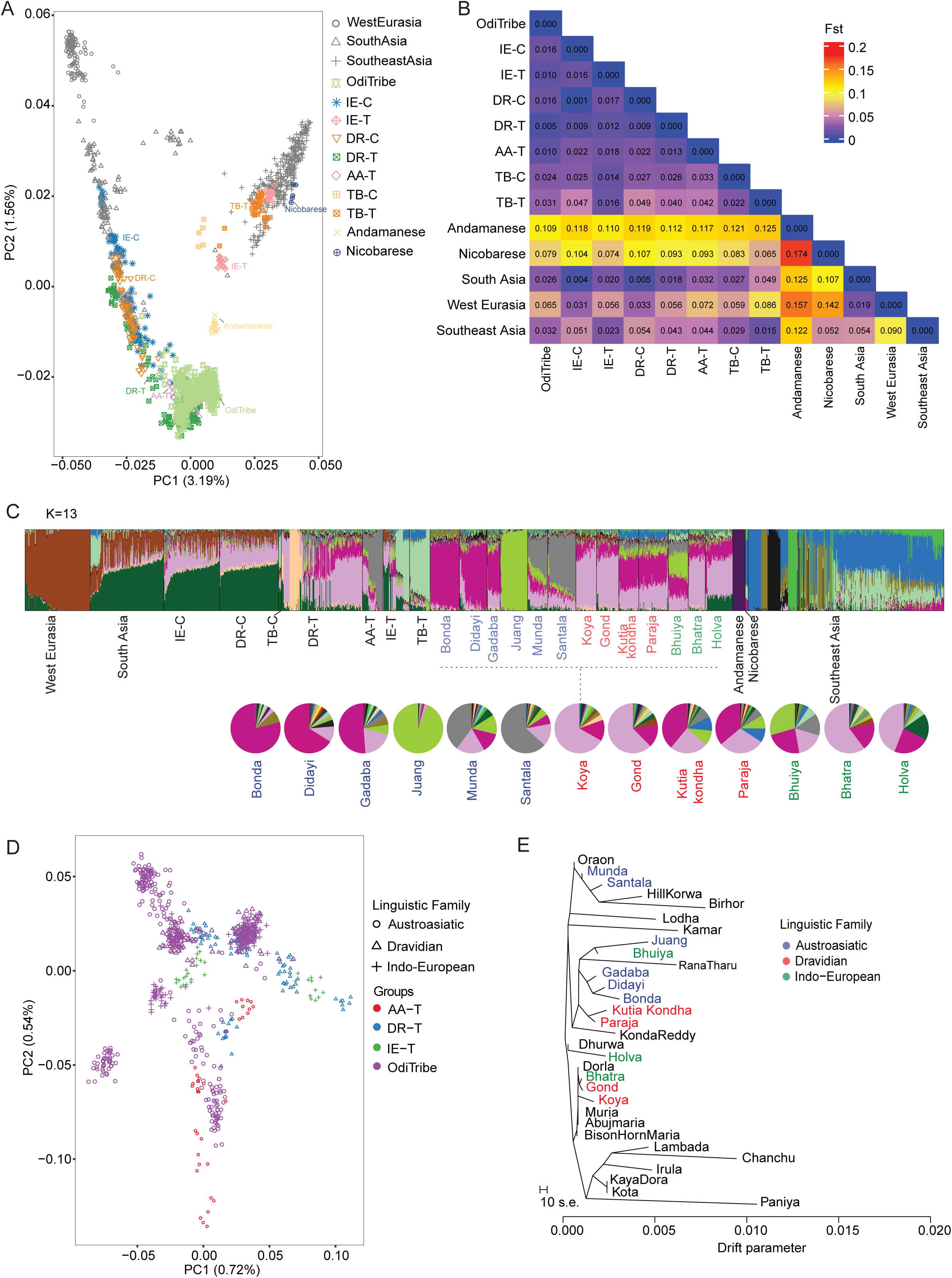
Genetic structure and ancestral composition of the OdiTribe. A. Principal component analysis of the OdiTribe (n = 515), West Eurasian, South Asian and Southeast Asian populations from the GenomeAsia 100K dataset (n = 1048). The Indian populations from the GenomeAsia 100K dataset are further classified into seven ethnolinguistic and social categories: Indo-European Caste (IE-C), Indo-European Tribe (IE-T), Dravidian Caste (DR-C), Dravidian Tribe (DR-T), Austroasiatic Tribe (AA-T), Tibeto-Burman Caste (TB-C) and Tibeto-Burman Tribe (TB-T). Shapes and colours denote the different populations and ethnolinguistic groups. B. Pairwise Wright’s Fst plot for OdiTribe and other population groups from the GenomeAsia 100K dataset. Blue color indicates a closer genetic relationship. C. Unsupervised admixture analysis of the individuals from OdiTribe and West Eurasian, South Asian and Southeast Asian populations from the GenomeAsia 100K dataset for K = 13. Bars are individuals, and colors denote ancestral components. The pie chart shows the mean admixture proportions in each OdiTribe group. D. Principal component analysis of OdiTribe (n=515), and DR-T, AA-T and IE-T groups from the GenomeAsia 100K dataset (n=187). Shapes denote the different linguistic families, and colours represent the OdiTribe and other tribal groups. E. TreeMix phylogeny of the OdiTribe groups (n = 515) with DR-T, AA-T and IE-T groups (n = 187) from the GenomeAsia 100K dataset, representing divergence patterns. The length of branches is proportional to the extent of population drift.

### Ancestral components of the OdiTribe

We used the unsupervised model-based clustering analysis implemented in the ADMIXTURE tool ^24^ for K values ranging from 2 to 18 to explore the ancestry of OdiTribe groups **(Supplementary Figure 3A-B)**. At a lower K (K=2 to K=3), the primary ancestral components resolved, separating West Eurasian, South Asian and Southeast Asian components. At higher K values, we found a finer substructure among the groups, corresponding to a combination of caste and linguistic families, i.e. IE and DR caste and tribal groups (ANI and ASI ancestries) and subsequently among AA (AAA ancestry) and TB (ATB ancestry) groups **(Supplementary Figure 3B)**. At K=9, the AAA ancestry component (grey colour) became evident, where Juang group appeared unadmixed compared to other AA groups indicating strong genetic drift and prolonged isolation (**Supplementary Figure 3B)**. The optimal number of K to explain the genetic differences was determined to be 13 based on the lowest cross-validation error value **(Supplementary Figure 3A)**. Among the AA-speaking OdiTribe groups, three components were identified. The AAA ancestry component (grey color), prominent among Santala (70%) and Munda (40%). The second was Bonda or the AAA-B component (fuchsia color), observed in Didayi (66%) and Gadaba (51%), followed by the third Juang or AAA-J component (lime-green color) **(Figure 2C)**. These AAA-B and the AAA-J components likely represent the population-specific drift rather than distinct ancestral sources. We also observed the presence of approximately 10% Southeast Asian ancestry in Bonda, Didayi and Gadaba and nearly 4% in Munda and Santala, consistent with ancient Southeast Asian-related gene flow across AA-speaking groups ^47^,^48^. In contrast, most of the DR and IE OdiTribe groups (with the exception of Bhuiya) were dominated by the ASI component (light purple), characteristic of the DR-T group **(Figure 2C)**. The individual tribal groups exhibited population-specific distinct components consistent with their linguistic affiliations and genetic diversity, as observed in the PCA **(Figure 2A)**.

### Shared genetic drift between OdiTribe and other Indian tribal populations

We used F3 outgroup statistics f3 (A; B, Yoruba), which measures the shared genetic drift between two populations since their divergence from the outgroup. The OdiTribe groups shared the highest level of drift among themselves and also with AA-T and DR-T groups **(Supplementary Figure 4A and Supplementary Table 4)**. Interestingly, the Juang (f3 =0.202175; Zscore = 61.529) and Bonda (f3 =0.202607; Zscore = 61.195) tribes exhibited more shared drift with the Nicobarese population **(Supplementary Figure 4B and 4C)**. This suggests an ancestral connection between mainland AA groups and island AA speakers. Didayi, Gadaba and Bhuiya showed substantial shared drift with Bonda and Juang, reflecting common ancestry and geographic continuity. In contrast, lower f3 values of OdiTribe groups with certain DR-T (Irula, Kota and Chanchu) and IE-T (Lambada, Rana Tharu) suggest limited gene flow **(Supplementary Table 4)**.

### Interplay of geography and language in shaping the OdiTribe genetic structure

We performed PCA of OdiTribe, DR-T, AA-T and IE-T to understand the genetic relationship in the regional context. Previous studies have mentioned that the Indian genetic landscape exhibits a stronger correlation with geographical regions than with linguistic families ^49^–^51^. In this study, the genetic clustering in the PCA reflects a complex interplay among language, geography and common ancestry **(Figure 2D and Supplementary Figure 5A)**. We found that the OdiTribe groups form a distinct but continuous cluster that partially overlaps with AA and DR tribes. This suggests that the OdiTribe shares ancestry with AA and DR tribal groups, consistent with their geographic proximity and shared demographic history in eastern and southern India. We also found internal substructure within the OdiTribe, which is likely shaped by endogamy and long-term isolation **(Figure 2D)**. When populations were grouped by geographical regions, a similar pattern was observed **(Supplementary Figure 5A)**. Tribal groups from eastern and central India clustered together, and those from southern India formed a neighboring continuum.

These observations were further corroborated by TreeMix ^25^ analysis. We observed two major clades corresponding to the AA and DR groups. In the AA clade, two sub-clades were observed **(Figure 2E)**. One sub-clade corresponding to Munda, Santala, Hill Korwa and Birhor, along with Oraon (DR group), indicating genetic continuity despite linguistic divisions. The second sub-clade consisted of Juang, Bhuiya (IE group), along with Rana Tharu (IE group), Bonda, Didayi and Gadaba **(Figure 2E)**. Notably, Kutia Kondha and Paraja (DR groups) also clustered within this clade, suggesting shared or common ancestry. Southern tribal groups such as Chanchu and Paniya were on longer branches, suggesting greater genetic drift and potentially smaller effective population sizes ^52^.

### Heterogeneity within OdiTribe populations

PCA of OdiTribe populations formed three major clusters corresponding to AA, DR and IE linguistic groups **(Figure 3A)**. Within the AA family, which is further subdivided into North and South Munda-speaking groups, we observed a clear differentiation **(Figure 3A)**. Juang, one of the South Munda-speaking groups, is genetically distinct from the other members of the family (Bonda, Didayi and Gadaba). Among the DR tribes, we observed two subclusters termed Dravidian group 1 (DR1) and Dravidian group 2 (DR2). DR1 cluster consisted of Paraja and Kutia Kondha, which clustered closely with neighbouring AA groups (Gadaba and Didayi). The second cluster, DR2, consisting of Gond and Koya, was positioned closer to IE groups such as Bhatra and Holva. These findings indicate possible historical gene flow or shared ancestry among the different linguistic families **(Figure 3A).** Subsequent principal components supported these observations, with Holva separating along PC2-PC3, showing separations within the IE-speaking groups (**Supplementary Figure 5B)**.

**Figure 3.**
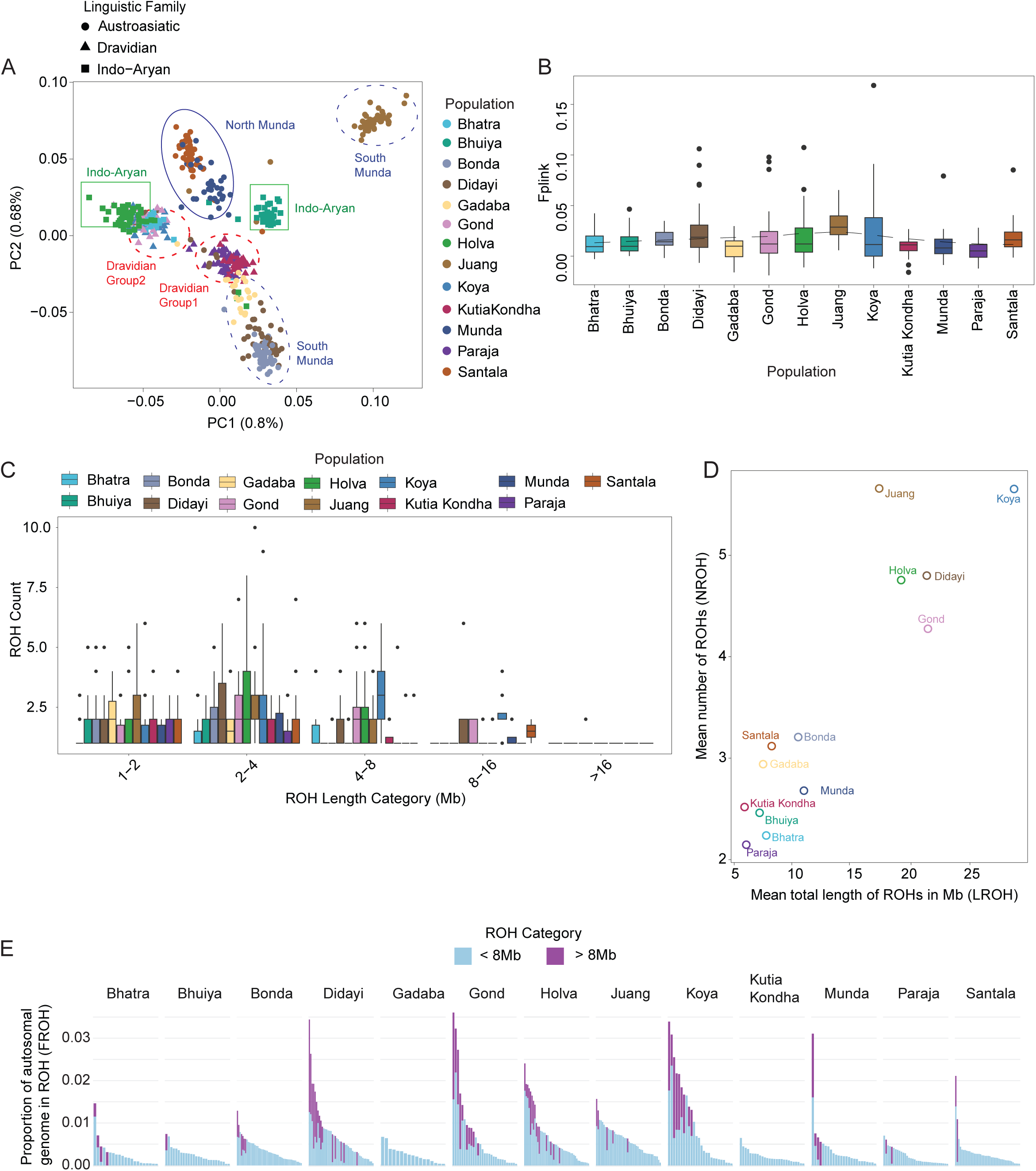
Inbreeding and runs of homozygosity among the OdiTribe groups. A. Principal component analysis on OdiTribe individuals. Shapes denote the different linguistic families, and colours indicate the different OdiTribe populations. B. Inbreeding coefficient (Fplink) across the OdiTribe groups. C. ROH segments across the populations. ROH regions spanning greater than 1 Mb genomic region are considered to be significant and were distributed into five classes based on length: class A (1 Mb–2 Mb), class B (2 Mb–4 Mb), class C (4 Mb–8 Mb), class D (8 Mb–16 Mb), and class E (>16 Mb). D. Relationship between the average number and the average sum of ROH segments per individual. E. The proportion of the genome covered by ROH regions below 8 Mb (1–8 Mb) and above 8 Mb.

We further measured the genetic divergence by Fst between the groups and built a neighbor-joining (NJ) tree **(Supplementary Figure 5C-D)**. The NJ tree mirrored the PCA clustering, further strengthening the linguistic and regional subcluster among the OdiTribe groups. Notably, the Juang tribe exhibited the highest branch length, indicating a greater genetic drift, followed by Holva, which also showed evidence of drift relative to other IE groups **(Supplementary Figure 5D)**.

Admixture analysis also captured fine-scale ancestral heterogeneity among the OdiTribe populations. K=2, identified two broad ancestry components corresponding to the AA and DR language families **(Supplementary Figure 6B)**. Subsequent values of K delineated components within the language groups. At K=5, we observed unique components corresponding to Bonda (red), Juang (green), Santala (purple), Koya (blue) and Holva (orange) **(Supplementary Figure 6B)**. The Dravidian-speaking Kutia Kondha and Paraja shared major ancestry components with Austroasiatic-speaking Bonda, Didayi and Gadaba, which is consistent with PCA and Fst analyses **(Supplementary Figure 6B)**. At K = 6, which yielded the lowest cross-validation error **(Supplementary Figure 6A)**, a novel ancestry component (yellow) emerged that was predominant in Kutia Kondha and Paraja, distinguishing them from both DR and AA groups **(Supplementary Figure 6B)**. This likely reflects population-specific genetic drift and isolation following earlier admixture, leading to the development of unique genetic signatures in these groups. We also observed Bhuiya to be admixed with varying degrees of AA components. These results collectively reveal fine-scale population substructure within OdiTribe, shaped by linguistic divergence, regional isolation, and varying degrees of historical gene flow.

### Inbreeding status and runs of homozygosity among OdiTribe

We used the inbreeding coefficient to assess the extent of cryptic relatedness and endogamy within the OdiTribe groups. Approximately 43% of the individuals exhibited an inbreeding coefficient greater than 0.0156, i.e. a kinship greater than that of second cousin marriage ^27^,^53^ **(Figure 3B, Supplementary Table 5)**. Among the groups, the Juang tribe showed the highest proportion of inbred individuals (89.3%), followed by Didayi (61.7%), Santala (50%) and Bonda (46%). In contrast, Paraja (84%), Gadaba (77.2%), Kutia Kondha (76.4%), and Munda (70.5%) had the majority of individuals with inbreeding coefficients below 0.0156, reflective of relatively low levels of inbreeding **(Supplementary Table 5)**.

Cryptic relatedness can also be measured with the cumulative length runs of homozygosity (ROH). ROH tracts are also known to be enriched for rare and deleterious variants ^27^. Following the classification scheme by Machha et al., ^17^, we grouped ROH segments into 5 classes: class A (1Mb - 2Mb), class B (2Mb - 4Mb), class C (4Mb - 8Mb), class D (8Mb - 16Mb), and class E (>16 Mb) **(Figure 3C)**. ROHs were detected in 429 individuals. Most individuals harbored multiple ROH segments in the shorter length categories (1Mb - 2Mb and 2Mb - 4Mb), with fewer long ROH segments (>8 Mb), indicating a predominance of distant rather than recent parental relatedness **(Figure 3C)**. 73 individuals (14.2%) had an ROH >8 Mb in length. The longest ROH of 25.04Mb was detected in an individual from the Didayi group. The total ROH length (LROH) and number of ROHs (NROH) varied substantially among populations **(Supplementary Figure 7A-B, Supplementary Table 6)**. Populations such as Juang, Koya, Didayi, and Holva exhibited both higher average LROH (17-28 Mb) and NROH (>4.5), consistent with consanguinity and pronounced endogamy **(Figure 3D)**. We also calculated the proportion of the autosomal genome occupied by the cumulative length of the smaller ROH tracts (FROH) **(Supplementary Figure 7C)**. While most individuals had a larger fraction of their genome in short ROH (< 8Mb), individuals from Didayi, Gond, Holva and Koya exhibited substantial coverage by long ROH (> 8MB) (**Figure 3E**). This indicates the presence of recent inbreeding in these groups. These findings highlight considerable heterogeneity in inbreeding and demographic history within the OdiTribe populations, with some groups showing signatures of both ancient and recent parental relatedness.

### Functional classification of the variants

The total SNVs identified in the OdiTribe dataset were annotated for variant types and their functional impact (See methods for details). Among known SNVs, 31.07% annotated to exonic regions, while this proportion was slightly lower, approximately 26% among novel SNVs **(Supplementary Figure 8A, Supplementary Table 7)**. The majority of the exonic variants were singletons (35%). Functional classification of the known exonic SNVs showed that 50% were non-synonymous, 0.13% were start-loss, 0.62% were stop-gain, and 0.05% were stop-loss variants. In comparison, novel SNVs exhibited a higher proportion of non-synonymous ∼60%, 0.1% were start-loss, 1.2% were stop-gain, and 0.12% were stop-loss variants **(Supplementary Figure 8B, Supplementary** Table **7)**. Further, the functional assessment of variants revealed that the singleton and rare variants had a higher proportion of high- and moderate-impact effects compared to common and uncommon variants (**Supplementary Table 7)**. In the rare category, 0.5% variants were predicted to have high impact, and 19.11% showed a moderate functional impact **(Supplementary Figure 8C, Supplementary Table 7)**. To assess clinical relevance, we cross-referenced the variants with the ClinVar ^41^, GWAS catalogue ^54^, and PharmGKB ^55^ databases. In total, 44,926 had prior evidence in ClinVar, 3,095 had previously been associated with GWAS traits, and 733 were reported in PharmGKB, highlighting the presence of variants with potential clinical and pharmacogenomic significance in the OdiTribe population **(Table 2)**.

**Table 2:** Clinical relevance of variants.

|  | **Common**<br>**(118344)** | **Uncommon**<br>**(65214)** | **Rare**<br>**(231625)** | **AC=1**<br>**(210413)** |
| --- | --- | --- | --- | --- |
| **ClinVar** | 14524 | 6226 | 14277 | 9899 |
| **GWAS** | 2350 | 342 | 293 | 110 |
| **PharmGKB** | 635 | 58 | 34 | 6 |

### High allele frequency of exonic variants in OdiTribe

To assess the genetic drift in the OdiTribe population, we compared the minor allele frequencies (MAF) of the SNVs with those of the Indian population from the GenomeAsia100K ^18^ dataset. Analysis of SNVs with predicted high and moderate functional effects shared between OdiTribe and GenomeAsia Indian populations uncovered a notable shift in MAF distribution. Specifically, 2,875 SNVs (9.73%) exhibited an increased MAF in OdiTribe compared to the Indian population **(Figure 4A)**. Notably, among SNVs that are classified as rare (MAF < 1%) in the GenomeAsia Indian population, 10.3% exhibited elevated allele frequencies in OdiTribe samples (MAF > 1%). Similarly, 7.7% of uncommon SNVs (1% < MAF < 5%) in the Indian population showed increased allele frequency in OdiTribe samples (MAF > 5%) **(Figure 4A)**.

**Figure 4:**
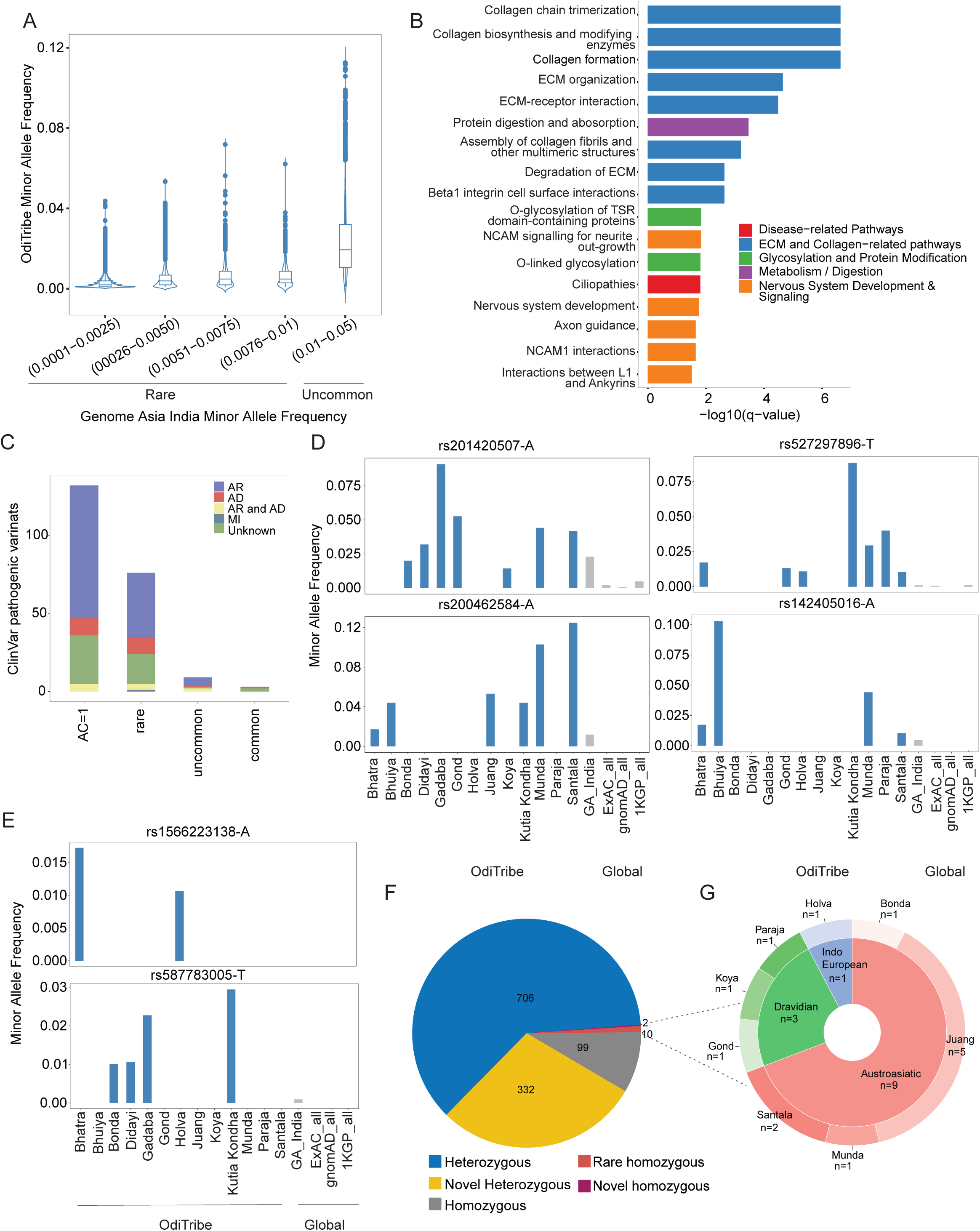
Variants of clinical importance in OdiTribe. A. Violin plot showing minor allele frequencies of variants from the GenomeAsia 100k Indian samples among OdiTribe. B. Pathways enriched for high-frequency variants in OdiTribe compared to the GenomeAsia 100k Indian samples. C. Distribution of variants in the OdiTribe dataset that are pathogenic in the ClinVar database. Autosomal-dominant (AD) or autosomal-recessive (AR) or mitochondrial inheritance (MI) other (unknown) classifications as per OMIM. D. Minor allele frequency plots of pathogenic variants in *DIO1* (top left), *GFI1B* (top left)*, MPZL2* (bottom left) *and PARK7* (bottom right) *genes* across OdiTribe groups and global populations. E. Carrier frequencies of ACMG actionable *BRCA2* (rs1566223138-A) and *BTD* (rs587783005-T) genes associated with dominant and recessive disorders compared with the global population cohorts. F. Proportion of heterozygous (known/novel) and homozygous (known/novel) HC-LOFs in the OdiTribe. G. Pie chart of novel homozygous HC-LOFs plotted by linguistic affiliation (inner circle) and OdiTribe group (outer circle).

To determine whether the genes harbouring these high-frequency SNVs are associated with specific biological pathways, we performed an over-representation analysis using the ConsensusPathDB database ^42^,^56^ A total of 2,875 variants with higher AF in the OdiTribe relative to the GenomeAsia Indian cohort were included in the study. These SNVs mapped to 2,289 genes. Of these, 1,533 genes were found to be enriched in at least one pathway. The over-representation analysis identified 17 significantly enriched biological pathways (q-value < 0.05, adjusted for multiple testing) (**Figure 4B, Supplementary Table 8**). The top enriched pathways were related to extracellular matrix (ECM) structure and remodeling, neural development and glycosylation processes. These findings suggest population-specific genetic adaptations that may influence structural integrity, neural function, and metabolic regulation. Notably, collagen gene families are important structural proteins and are known for their stronger negative selection in different ethnic groups ^57^. Neural development and cell adhesion pathways, including NCAM1 signaling and axon guidance, suggest population-specific variants may influence cognitive traits, neurodevelopment and susceptibility to neurological conditions ^58^,^59^.

### OdiTribe-specific novel variants and their functional impact

Among the novel non-singleton SNVs (n=34,667), 84.9% (n=29,440) were OdiTribe specific, 14.8% (n=5,139) were shared by other Indian populations (GenomeAsia Indian cohort), and 0.25% (n=88) were shared by other Asian populations (GenomeAsia Asian cohort) **(Supplementary Figure 8D)**. The distribution of the OdiTribe-specific novel SNVs reflects the genetic subcluster within the tribal groups. Populations showing distinct clustering in PCA and Fst analyses also exhibited unique and shared novel variant patterns, suggesting that historical isolation and limited gene flow have contributed to the accumulation of group-specific variants. Specifically, Juang, the least admixed group, harboured the highest number of unique novel variants (n=2,733), followed by Bonda (n=2,182) and Holva (n=2,115) **(Supplementary Figure 8E)**. The highest number of shared novel variants was observed between Didayi and Bonda (n = 1,165), both belonging to the AA family, consistent with their close genetic proximity. Similarly, Paraja and Kutia Kondha (DR groups) shared 841 novel variants, aligning with their clustering in PCA and lower pairwise Fst values. Interestingly, despite belonging to different linguistic groups, Juang and Bhuiya shared 621 novel variants, suggesting historical admixture or shared ancestral variation. Overall, the distribution of novel SNVs within the OdiTribe mirrors the population substructure, reflecting the combined effects of linguistic affiliation, geographic proximity, and genetic drift.

Given that we observed an excess of unique and shared OdiTribe-specific novel variants, we evaluated the functional effect of these variants and their association with the diseases using different approaches (See methods for details). A total of 43 potentially deleterious novel variants in 42 genes were identified in the OdiTribe dataset **(Supplementary Table 9)**. All these variants were present in exonic regions with non-synonymous variants. Importantly, 8 novel potentially deleterious variants in 7 genes showed homozygosity in 11 individuals **(Supplementary Table 9).**

### Pathogenic variants and probable clinical implications

Identified OdiTribe variants were annotated against the ClinVar database ^41^. A total of 220 SNVs in 193 genes were identified as pathogenic/likely-pathogenic (P/LP) and further examined for OMIM inheritance patterns. We observed SNVs with an autosomal recessive mode of inheritance to be prominent in OdiTribe populations **(Figure 4C).** We identified a total of 44.6% (n=230) individuals harbouring at least one P/LP variant identified in 131 recessive condition genes ^46^ and 10.8% (n=56) individuals were carriers for multiple recessive conditions. Notably, ∼24% of SNVs had unknown inheritance patterns, indicating gaps in current genetic knowledge. More than 60% (n=132) of pathogenic SNVs were singletons, which suggests the potential clinical relevance of the OdiTribe population **(Figure 4C)**. Next, we explored pathogenic variants that are highly prevalent (MAF > 0.01) in the OdiTribe cohort but rare (MAF < 0.01) in the global population gnomAD ^39^, ExAC ^60^, 1000 Genomes ^40^ databases. Six pathogenic mutations in six genes were identified at high frequencies in the OdiTribe population compared to the global population (**Supplementary Table 10)**. Notably, a splice donor variant in the *HBB* gene (rs33915217-G) and an intronic variant in the *HBD* gene (rs549964658-A) are associated with beta-thalassemia and delta-thalassemia ^61^,^62^ respectively were identified. Both the variants are rare in all super populations from gnomAD ^39^, ExAC ^60^ and 1000 Genomes ^40^ datasets and but not rare (MAF > 0.01) in the GenomeAsia India population ^18^, suggesting high prevalence of these variants in the South Asian population **(Supplementary Figure 9A)**. When we classified the GenomeAsia India population into tribe and caste groups we observed a higher minor allele frequency in OdiTribe groups **(Supplementary Figure 9A)**. The minor allele frequency was 0.08 and 0.17 for rs33915217-G and rs549964658-A mutations in the Paraja and Holva groups, respectively **(Supplementary Figure 9A)**. Further, we analysed biochemical parameters among individuals carrying the *HBB* (rs33915217-G) and *HBD* (rs549964658-A) mutations. Interestingly, individuals with a heterozygous mutation in *HBB* gene showed significantly lower mean corpuscular hemoglobin concentration (MCHC) (p < 0.01), and red cell distribution width (RDW) was significantly higher compared to normal individuals (p < 0.001) **(Supplementary Figure 9B)** and individuals with homozygous mutation in *HBD* gene had elevated levels of RBC count (p < 0.05) **(Supplementary Figure 9C)**. This indicates the potential impact of this mutation on hematological parameters and following it further for genetic counselling in the respective populations. Additionally, the other four pathogenic missense mutations in *DIO1* (rs201420507-A), *GFI1B* (rs527297896-T), *MPZL2* (rs200462584-A) and *PARK7* (rs142405016-A) are reported to be associated with thyroid hormone metabolism ^63^, platelet-type bleeding disorder ^64^, hearing loss ^65^ and early-onset Parkinson’s disease ^66^ were identified **(Figure 4D)**. Each of these variants is at a high frequency in at least one of the OdiTribe groups but rare in the super-populations and other Indian tribe and caste populations **(Figure 4D, Supplementary Figure 9D)**. Overall, these observations indicate the importance of including an ethnically diverse populations in human genetic studies, as the rare variant threshold is used to determine the pathogenicity of the variants in clinical genomic studies ^1^,^67^.

Furthermore, we also examined the distribution of missense variants in the 81 medically actionable genes recommended by ACMG (v3.2) ^45^. A total of 326 exonic variants (known=306, novel=20) consisting of 322 missense, 2 stop gain and 2 unknown variants were distributed across 56 actionable genes **(Supplementary Table 11).** The variant burden was found to be highest in recognized disease-associated genes *APOB* (n=31), *BRCA2* (n=23), *BRCA1* (n=20), *GAA* (n=18) and *APC* (n=16) **(Supplementary Figure 10A)**. Further, these variants are classified based on clinical significance according to the ClinVar database ^41^ **(Supplementary Figure 10B)**. Among these 3 pathogenic or likely pathogenic (P/LP) variants, notably *BTD* (2 variants) and *BRCA2* (1 variant) were identified. Interestingly, both variants in the *BRCA2* and *BTD* gene (rs1566223138-A, rs587783005-T, respectively) are completely absent or ultra-rare in the global populations, highlighting their potential clinical and population-specific relevance **(Figure 4E, Supplementary Figure 10C-D)**. Additionally, our analysis also uncovered 20 novel variants specific to the OdiTribe population distributed across 12 ACMG-recommended genes. Among these, *APOB* harbored the highest number of novel variants (n = 5), followed by *BRCA2* (n = 2) and GAA (n = 2) (**Supplementary Table 11).** These findings highlight the potential clinical relevance of the OdiTribe population and the necessity of population-scale genomic studies in these underrepresented ethnic groups.

### Homozygous predicted loss-of-function variants

Loss-of-function (LoF) variants may affect both alleles of a gene, and studying the phenotypes associated with these genes can enhance our understanding of their function. The prevalence of endogamy and consanguineous marriages increases the likelihood of offspring inheriting homozygous loss-of-function mutations ^68^. To understand the consequences of variants with LoF in our dataset, we annotated the variants for predicted loss-of-function mutations (pLOFs) using the LOFTEE tool ^32^. We identified 2295 LoF variants, of which 610 (26.5%) are novel **(Supplementary Figure 10D)**. Furthermore, we identified 1041 high-confidence LoF (HC-LoF) variants in 956 genes across the OdiTribe dataset, of which 520 (49.9%) were singletons (**Supplementary Table 12)**. A major proportion of variants were splice donor and stop gain variants for HC-LoF and low-confidence LoF (LC-LoF) groups, respectively **(Supplementary Figure 10E)**. Next, we focused only on HC-pLoF; among these, 332 HC-LoFs were novel, and 709 were known variants **(Figure 4F)**. Most of these were heterozygous variants and identified 111 homozygous HC-pLoF variants in 110 genes in our OdiTribe dataset and compared them with the homozygous pLoF lists from GenomeAsia ^18^ and gnomAD ^39^. We identified a total of 75 genes with homozygous pLoFs specific to the OdiTribe dataset **(Supplementary Table 12)**. Since common pLoF variants are less likely either to have a functional effect or to be subject to purifying selection ^27^,^69^, we first analysed the number of high-confidence homozygous pLoF variants with an allele frequency lower than 1% in the OdiTribe dataset. We uncovered 12 rare homozygous HC-pLoFs in 12 genes **(Figure 4G and Supplementary Table 12)**. These homozygous pLoFs were observed in 13 individuals (2.52%), 11 individuals with single HC-pLoFs and one individual with two HC-pLoFs **(Figure 4G)**. Specifically, the Juang tribe group harboured the highest HC-pLoFs (n=5) compared to other groups, out of which 2 pLoF variants in 2 genes were novel. Both the splice variant HC-pLoFs in *ISM2* (chr1:46938138-C>G) and *CYP4A11* (chr14:77484921-T>A) gene were predicted as moderate pathogenic and supporting pathogenic with a CADD score ^37^ of 34 and 27.6, respectively. Both the genes *CYP4A11* and *ISM2* are associated with blood pressure regulation and hypertension ^70^,^71^ indicating loss of function variant in *CYP4A11* gene may reduce the enzymès ability to metabolize fatty acids which can change the blood pressure regulation and homozygous loss of function variant in *ISM2* gene can destabilize the protein structure and leads to change in protein expression ^70^,^71^ Both variants are found in 10 individuals, predominantly present in the Juang tribal group (**Supplementary Table 12)**.

Homozygous pLoFs with allele frequency higher than 1% might suggest positive selection or the pseudogenization effect of genes ^72^. We extracted common homozygous HC-pLoFs from the OdiTribe cohort and identified 99 common homozygous HC-pLoFs in 98 genes **(Supplementary Table 12)**. We then compared our list with the previously reported homozygous pLoFs from gnomAD ^39^ and ExAC ^60^. We identified 89 genes with common homozygous HC-pLoFs that were also present in the list of ExAC/gnomAD, and 10 genes are unique to our OdiTribe dataset **(Supplementary Table 12)**. Overall, our genomic analysis of indigenous and highly endogamous tribal groups from Odisha identified the importance of deep genomic profiling of these isolated populations to build a comprehensive catalogue of disease-impacting genomic variations.

## Discussion

The demographic history of the Indian subcontinent is remarkably complex and diverse, which is shaped by multiple waves of migration and historical events, diverse environmental exposures, cultural practices and the practice of endogamy among ethnic groups ^52^. Many present-day Indian populations’ origin can be traced back to a limited number of founding groups maintaining distinct genetic identities through endogamy.

In this study, we provide one of the most extensive catalogues to date of genetic diversity in indigenous tribal populations represented in one of the most diverse tribal group-occupied regions of India. Using high-depth whole-exome sequencing of 515 individuals representing 13 tribal groups, we uncovered a unique and previously underrepresented collection of genetic variation in eastern India. We identified 6,25,596 high-confidence SNVs, which included 2,10,413 singletons (33.6%), and 34,667 novel non-singleton variants (5.54), that are not observed in global datasets. We also observed the predominance of rare variants in our dataset. Tribes such as Gadaba, Munda and Bhuiya exhibited both elevated counts of SNVs and singletons, whereas Juang and Bonda showed reduced diversity likely due to stronger drift and historical bottlenecks. The linguistic stratification of genetic variation revealed that AA groups harbored both fewer variants and singletons than DR and IE groups, suggesting a lower genetic diversity among AA-speaking groups. While most of the common variants (MAF > 0.05) were shared across the three linguistic families, rare variants exhibited limited overlap, indicating both shared ancestry and subsequent differentiation.

Our analysis of the population structure identified a complex genetic architecture influenced by geography, language, and endogamy. Principal component analysis using the global dataset placed the OdiTribe groups in proximity to other DR and AA tribal groups of central and eastern India, consistent with their geographic and ethnolinguistic affiliations. The observed position of OdiTribe at the distal end of the South Asian cline and its differentiation from the caste groups emphasise strong continuity with geographically proximate tribal groups and limited gene flow with the caste groups, who display more admixed profiles due to historical gene flow with Indo-European and Central Asian groups ^73^. ADMIXTURE analysis revealed complex ancestral composition and fine-scale genetic structure among the OdiTribe groups. We identified three AA ancestry components among the Munda-speaking groups which might be due to population-specific drift and prolonged isolation. The presence of a minor fraction of Southeast Asian ancestry (∼10%) observed in Bonda, Didayi, and Gadaba is consistent with ancient gene flow between island and mainland AA groups along the Bay of Bengal corridor ^48^,^49^. The DR-speaking groups, Paraja and Kutia Kondha, exhibited shared ancestry with neighboring AA groups, indicating regional gene flow. These observations indicate that the genetic diversity of the OdiTribe population is shaped by admixture and subsequent genetic drift, and founder effect, leading to population-specific components.

PCA with OdiTribe in combination with other Indian tribes (DR-T, AA-T and IE-T) supported a multifaceted relationship among language, geography, and common ancestry. We found that the OdiTribe groups form distinct but continuous clusters that overlap with both AA and DR tribes and are distant from the IE tribes. The OdiTribe groups occupied a genetic space overlapping primarily with central and eastern tribal groups and adjacent to southern tribes, suggesting a deep ancestral connection of OdiTribe groups with AA and DR tribe groups, consistent with their geographic proximity and shared demographic history in eastern and southern India.

Local substructure within OdiTribe was also seen in the PCA, where North and South Munda-speaking groups separated into distinct clusters, indicative of ancestral divergence, which may be further enhanced by geographic isolation. We also observed two subclusters among the DR-speaking OdiTribe groups, which was further corroborated by measuring genetic divergence using Fst, indicating possible historical gene flow or shared ancestry among the groups from different linguistic families.

Inbreeding coefficient and runs of homozygosity analyses revealed differences in marriage practices among the groups. Approximately 43% of the OdiTribe individuals exhibited inbreeding coefficients higher than the threshold for second cousins, suggesting persistent endogamy and localized consanguinity. Groups such as Juang, Didayi and Holva exhibited long ROH segments (>8 Mb), which indicates recent parental relatedness and small effective population sizes. In contrast, Paraja and Munda displayed shorter ROH segments, indicating older founder events. This heterogeneity in the ROH segments highlights variable demographic histories, consistent with multiple founder events, geographic isolation, and the practice of endogamy among South Asia groups Functional annotation of variants revealed a rich catalogue of both known and novel variants in the OdiTribe dataset. Comparing minor allele frequencies between OdiTribe and the Indian population from the GenomeAsia dataset revealed a significant shift in the frequency of functionally important variants in the OdiTribe dataset. Approximately 10% of variants classified as rare (MAF < 0.01) in other Indian populations were observed at higher frequencies within OdiTribe. Pathway enrichment analysis of genes harboring high frequency variants revealed pathways related to extracellular matrix (ECM) structure and remodeling, neural development and glycosylation processes.

Additionally, 220 pathogenic or likely pathogenic variants across 193 genes were identified. The predominance of variants with a recessive mode of inheritance and a high proportion of singletons among pathogenic variants reflect the impact of endogamy and consanguinity on the accumulation of deleterious alleles. Notably, 44.6% of the OdiTribe individuals carried at least one pathogenic variant associated with a recessive inheritance pattern and 10.8% were carriers of multiple recessive alleles. This high carrier load highlights the potential risk for recessive disorders in small, endogamous groups and also underscores the need for population-specific screening strategies for recessive diseases.

Previous reports have documented a high prevalence of infectious diseases such as malaria, tuberculosis, as well as genetic disorders like G6PD deficiency, sickle cell anemia, and thalassemia among the tribal groups (ICMR Bulletin, October 2003) ^16^. Although no pathogenic variants related to sickle-cell anemia or G6PD deficiency were identified in the OdiTribe dataset, we detected two known pathogenic variants in *HBB* (rs33915217-G) and *HBD* (rs549964658-A) genes, which are associated with beta and delta thalassemia, respectively. These variants occurred at higher frequencies in Paraja (AF□=□0.08) and Holva (AF□=□0.17), respectively. Moreover, we observed altered hematological parameters among carriers, further demonstrating the phenotypic impact of these variants and underscoring the need for genetic counselling and awareness within these affected communities.

In addition, we found missense pathogenic variants in *DIO1* (rs201420507-A), *GFI1B* (rs527297896-T), *MPZL2* (rs200462584-A) and *PARK7* (rs142405016-A) mutations with increased abundance in OdiTribe groups. Notably, the variant in the *MPZL2* gene was observed at high allele frequency (AF=0.12) in Santala. Loss-of-function mutations in *MPZL2* are known to cause autosomal recessive nonsyndromic hearing loss characterized by early-onset moderate hearing impairment ^65^. A mutation in *PARK7* was identified in the Bhuiya (AF=0.10). The gene encodes the DJ-1 protein, which plays a critical role in mitochondrial homeostasis and oxidative stress response ^74^. Mutations in this gene have been implicated in the early onset of autosomal recessive Parkinson’s disease ^66^. Although the overall prevalence of this disease is globally low, the relatively high allele frequency observed in our dataset highlights the undiagnosed cases within isolated endogamous groups and underscores the importance of genetic screening and awareness within these groups.

In addition, we also identified several exonic variants across 56 ACMG-recommended actionable genes, including *APOB*, *BRCA1/2*, *GAA* and *APC* are the top frequently mutated genes. The high mutation burden observed in BRCA1/2, including one pathogenic variant, suggests potential predisposition to hereditary cancer syndromes within the OdiTribe population. Furthermore, the discovery of 20 novel OdiTribe-specific variants highlights the lack of representation of indigenous South Asian populations in global medical genomics resources.

In endogamous populations, the probability of homozygous LoF alleles increases due to consanguinity and genetic drift, thus offering a unique opportunity to study the phenotypic consequences of gene disruption ^68^. In the OdiTribe dataset, we observed a significant number of LoF variants. Notably, several homozygous HC-LoF variants were found in genes associated with interferon signaling and protein ubiquitination processes. Notably, a homozygous HC-LoF variant in the *TRIM34* gene was identified in a single individual from the Bonda group. Evidence suggests that loss of function of *TRIM34* leads to increased susceptibility to certain diseases such as colitis associated colorectal cancer and influenza A infection ^75^,^76^. Overall, our comparative analysis across language groups indicates the tribal groups belonging to Austroasiatic language families including Bonda, Didayi and Juang harbor higher number of homozygous HC-LoF variants compared to other OdiTribe groups underscoring the impact of long-term isolation and demographic history on functional genetic variation.

In summary, our study presents an exome-wide characterization of population-specific variants, offering comprehensive insights into the genetic differentiation and ancestral composition among the tribal population of Odisha. Disease-causing genetic variants which are rare in global populations may occur at higher frequencies within such isolated endogamous groups as evident in the study. Since the access to medical resources is limited among the tribal groups these variants and the respective underlying disorders mostly remain undiagnosed. Large-scale genetics studies such as ours can form the basis for screening of hereditary pathogenic variants, offering a scope for targeted healthcare interventions.

## Declaration of interests

The authors have no conflict of interest to declare.

## ILS Tribal Flagship Consortium

Debasis Dash, Gulam Hussain Syed, Rupesh Dash, Satish Devadas, Shantibhusan Senapati, Soma Chattopadhyay, Sunil Kumar Raghav, Tushar Kant Beuria, V.Arun Nagraj

## Acknowledgements

We acknowledge late Dr. Ajay Parida for initiating the tribal flagship project and Dr. Debasish Dash for his support throughout the project. We sincerely thank all the participants who volunteered for this study. We are grateful to the social workers and Ministry of Tribal Affairs for their invaluable assistance. We also thank Dr. Mitali Mukherji, Dr. Swarkar Sharma, Dr. Sabarinathan Radhakrishnan, Dr. Vinod Scaria and Dr. Nidhan Biswas for their valuable scientific discussions and suggestions. We extend our appreciation to the ILS Flagship team for coordinating the sample collection, the ILS Next Generation Sequencing facility and the ILS High Performance Computing facility for their technical support. We also thank Mr. Shaktiprasad Mishra for his help with tribal literature review. Finally, we acknowledge the financial assistance received as student fellowship from the Indian Council of Medical Research (ICMR).

## Author Contributions

SKR, PP and ILS Flagship Consortium conceived the study. SD, DJ and AG performed sequencing and data analysis along with VM. SJ and SM conducted field visits and carried out anthropological survey and sample collection. VKB, SJ and MS conducted whole exome library preparation. SM and RM curated the metadata. SD, DJ and AG wrote the original draft. SKR, AB and BG reviewed and edited the manuscript.

## Web Resources

dbNSFP – https://sites.google.com/site/jpopgen/dbNSFP

gnomAD – https://gnomad.broadinstitute.org

1000 Genomes Project – https://www.internationalgenome.org

ClinVar – https://www.ncbi.nlm.nih.gov/clinvar

GenomeAsia – https://genomeasia100k.org

ConsensusPathDB – http://cpdb.molgen.mpg.de

dbSNP – https://www.ncbi.nlm.nih.gov/snp

OMIM (Online Mendelian Inheritance in Man) – https://www.omim.org

## Data and Code availability

The analyzed dataset presented in this study is available from the corresponding author upon reasonable request.

## Supplementary Figure Legends

**Supplementary Figure 1: Sample-based quality metrics**

A. Mean target coverage for all the OdiTribe samples (n=634) (left) and percentage of the target region covered across the different coverages. The horizontal line marks 95% of the target region covered (right).

B. Percentage of OdiTribe individuals (n=634) in each linguistic family: Austroasiatic (AA), Dravidian (DR), and Indo-European (IE).

C. Number of males and females in the OdiTribe dataset (n=634) (left) and age distribution across the genders (right).

D. Mean target coverage for 515 the OdiTribe samples after QC (left) and percentage of the target region covered across the different coverages. The horizontal line marks 95% of the target region covered (right).

E. Number of individuals (n=515) across the thirteen OdiTribe groups after quality control. Groups are colored according to their linguistic family.

F. Distribution of total SNVs (right) and singletons (left) across the linguistic families.

**Supplementary Figure 2. Population structure analysis**

A. Principal component analysis of OdiTribe (n=515), West Eurasia, South Asia and Southeast Asian populations from the GenomeAsia 100K dataset (n=1048). The Indian populations from the GenomeAsia 100K dataset are further classified into seven ethnolinguistic and social categories: Indo-European Caste (IE-C), Indo-European Tribe (IE-T), Dravidian Caste (DR-C), Dravidian Tribe (DR-T), Austroasiatic Tribe (AA-T), Tibeto-Burman Caste (TB-C) and Tibeto-Burman Tribe (TB-T). Shapes and colors denote the different populations and ethnolinguistic groups. Plot for the second and third principal components are shown. PC3 separates Andamanese from the other populations.

B. Principal component analysis of OdiTribe (n=515), West Eurasia, South Asia and Southeast Asian populations from the GenomeAsia 100K dataset (n=1048). Shapes indicate the different population groups, and colors represent the various Indian geographical regions the ethnolinguistic groups belong to. Plot for the first and second principal components are shown.

**Supplementary Figure 3. Admixture analysis of OdiTribe in a global context**

A. Cross-validation error for OdiTribe (n=515), West Eurasia, South Asia and Southeast Asian populations from the GenomeAsia 100K dataset (n=1048). K=18 resulted in the lowest CV error.

B. Unsupervised admixture analysis for K=2-18. Bars are individuals, and colors denote ancestral components

**Supplementary Figure 4. Shared genetic drift between the OdiTribe groups and other Indian tribes**

A. Outgroup **f**_3_ test of the form f_3_ (OdiTribe group, X; Yoruba), where X represents other tribal groups. Error bars indicate jackknife-derived standard errors, highlighting the relative allele sharing between the study populations and tribal groups.

B. Outgroup **f**_3_ plot of Juang of the form (Juang, X; Yoruba), where X represents other tribal groups. Error bars indicate jackknife-derived standard errors, highlighting the relative allele sharing between the study populations and tribal groups.

C. Outgroup **f**_3_ plot of Bonda of the form (Bonda, X; Yoruba), where X represents other tribal groups. Error bars indicate jackknife-derived standard errors, highlighting the relative allele sharing between the study populations and tribal groups.

**Supplementary Figure 5. Substructure within the OdiTribe groups**

A. Principal component analysis of OdiTribe (n=515), DR‐T, AA‐T and IE‐T groups from the GenomeAsia 100K dataset (n=187). Shapes denote the different geographical regions where each group belongs, and colors represent the other tribal groups. Plots for the first and second principal components are shown.

B. Principal‐component analysis on OdiTribe groups. The second and third principal components are shown. Shapes are the different linguistic families, and colors are the tribal groups.

C. Pairwise Wright’s Fst plot for the OdiTribe groups. Blue color indicates a closer genetic relationship.

D. Neighbor‐joining tree based on Wright’s Fst values. The longer the branch, the higher the genetic differentiation.

**Supplementary Figure 6. Admixture analysis of OdiTribe individuals**

A. Cross‐validation error for OdiTribe individuals. K=6 resulted in the lowest CV error.

B. Unsupervised admixture analysis for K=2‐8. Bars are individuals, and colors denote ancestral components

**Supplementary Figure 7. Runs of homozygosity analysis across the OdiTribe groups**

A. Distribution of number of ROH segments (NROH) across the OdiTribe groups

B. Distribution of total length of ROH segments (LROH) across the OdiTribe groups

C. Distribution of proportion of autosomal genome in ROH (FROH) across the OdiTribe groups

**Supplementary Figure 8. Functional annotation of the variants**

A. Percentage of known, novel variants and singletons (AC=1) classified according to the genomic region.

B. Percentage of known, novel variants and singletons (AC=1) classified according to the variant type

C. Percentage of variants predicted to have functional impact across rare (MAF <0.01), uncommon (0.01 > MAF < 0.05) and common (MAF > 0.05) variant categories and singletons (AC=1).

D. Percent shared novel variants (AC > 1) compared between OdiTribe, the Indian population and other Asian populations from the GenomeAsia100K dataset.

E. Distribution of unique and shared novel variants across the OdiTribe groups.

**Supplementary Figure 9. Clinically important 1179 pathogenic variants**

A. Minor allele frequency plot of pathogenic variants in the *HBB* (rs3391527‐G, top) and *HBD* (rs549964658‐A, bottom) genes across OdiTribe groups and global population cohorts (left) and other Indian tribe and caste population (right).

B. Biochemical plot for RDW.CV and MCHC levels for the rs3391527‐G variant for the OdiTribe dataset.

C. Biochemical plot for RBC count for thers549964658‐A variant for the OdiTribe dataset.

D. Minor allele frequency plots of pathogenic variants in *DIO1* (top left), *GFI1B* (top left), *MPZL2* (bottom left) and *PARK7* (bottom right) genes across OdiTribe groups and other Indian tribe and caste population.

**Supplementary Figure 10. Pathogenic variants in ACMG actionable genes and distribution of predicted loss‐of‐function (pLoF) genes**

A. Proportion of known and novel variants detected across the ACMG actionable genes

B. Proportion of variants across the ACMG actionable genes grouped by their clinical significance according to the ClinVar database.

C. Minor allele frequency plot of pathogenic variant in ACMG recommended actionable *BTD* gene (rs587783005‐T) across OdiTribe groups and other Indian tribe and caste population.

D. Minor allele frequency plot of pathogenic variants in the *BTD* (rs13078881‐C) gene across OdiTribe groups and global population cohorts (left) and other Indian tribe and caste population (right).

E. Number of known and novel variants across low confidence (LC‐pLoF) and high confidence (HC‐pLoF) putative loss‐of‐function categories.

F. Number of LC‐pLoF and HC‐pLoF across the MAF categories (rare, uncommon and common) when grouped by the variant type. AC=1 denotes singletons.

## Legend for Supplementary Tables

Supplementary Table 1: Total SNV and singleton statistics for each population group

Supplementary Table 2: Total SNV and singleton statistics for each language group

Supplementary Table 3: Distribution of rare (MAF < 0.05) and common (MAF > 0.05) variants across each language group

Supplementary Table 4: F3 outgroup analysis

Supplementary Table 5: Inbreeding coefficient calculated for each individual

Supplementary Table 6: ROH summary for each individual

Supplementary Table 7: Functional annotation of variants

Supplementary Table 8: List of pathways enriched for genes with high minor allele frequency in OdiTribe compare other Indian population

Supplementary Table 9: Functional impact of novel variants

Supplementary Table 10: List of pathogenic variants in OdiTribe

Supplementary Table 11: List of exonic variants in ACMG recommended actionable genes

Supplementary Table 12: List of high confidence loss of function (HC‐LoF) variants in OdiTribe

## Notes

### Competing Interest Statement

The authors have declared no competing interest.

### Funding Statement

The project was funded by the Department of Biotechnology, Ministry of Science and Technology, Government of India (Grant id: BT/ILS/Flagship/2019).

### Author Declarations

The Institutional Ethics Committee of the Institution of Life Sciences gave ethical approval for this work (IEC no: 87/HEC/19).

