## Supplementary figures and images for "Fine-scale genetic structure and population-specific clinically relevant variants in the indigenous tribal populations of India"

### Supplementary Figure 1

#### Supplementary Figure 1

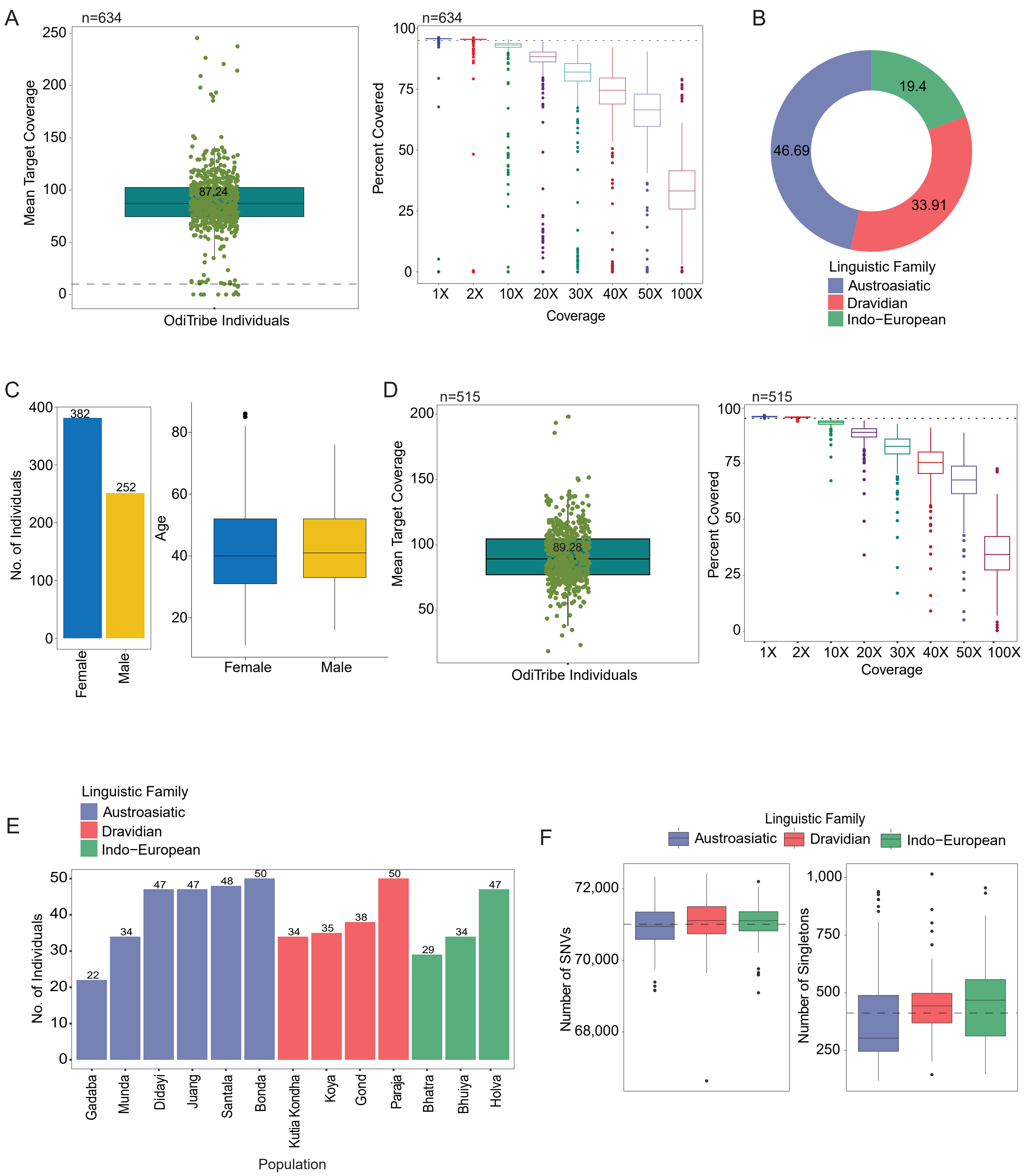

### Supplementary Figure 2

#### Supplementary Figure 2

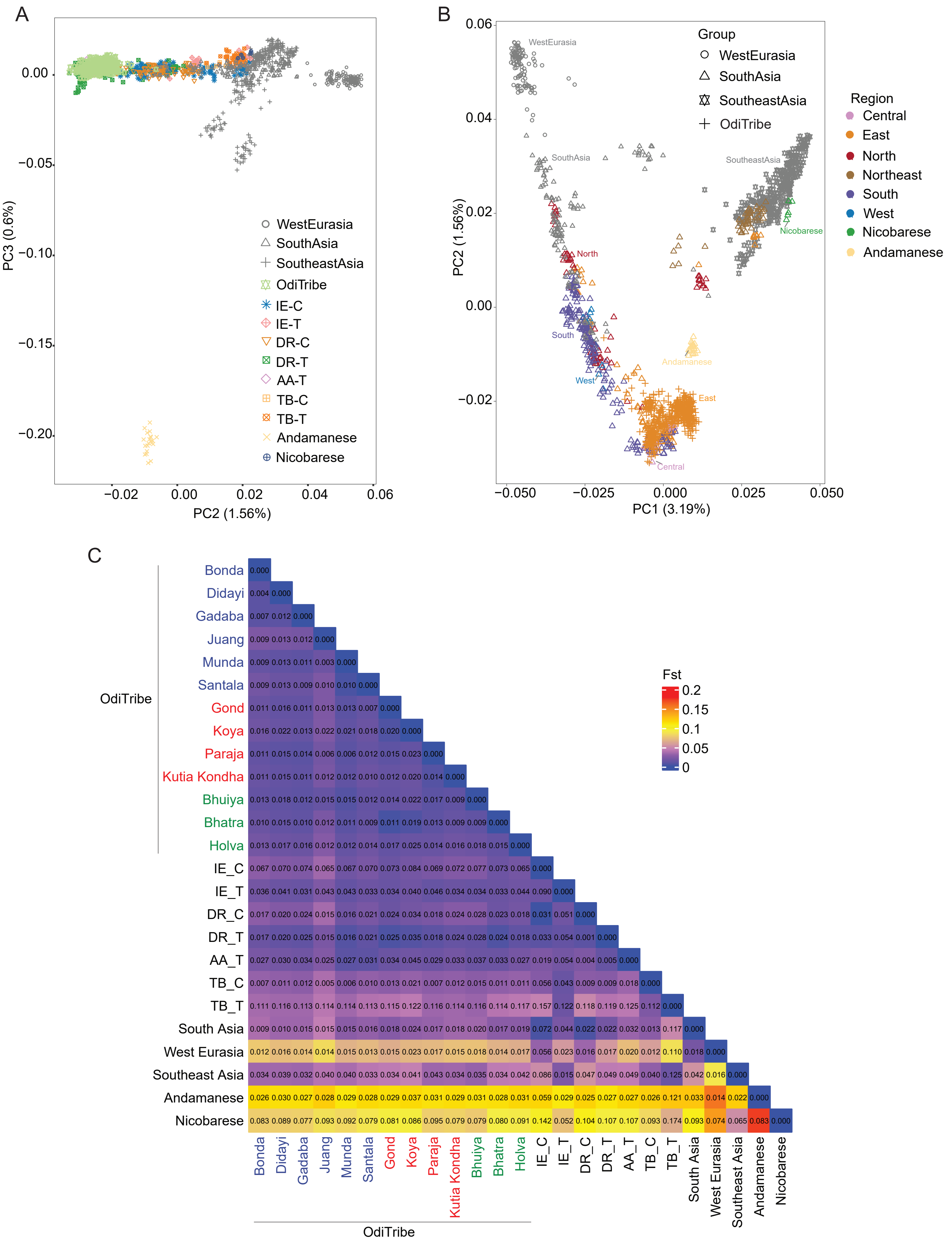

### Supplementary Figure 3

#### Supplementary Figure 3

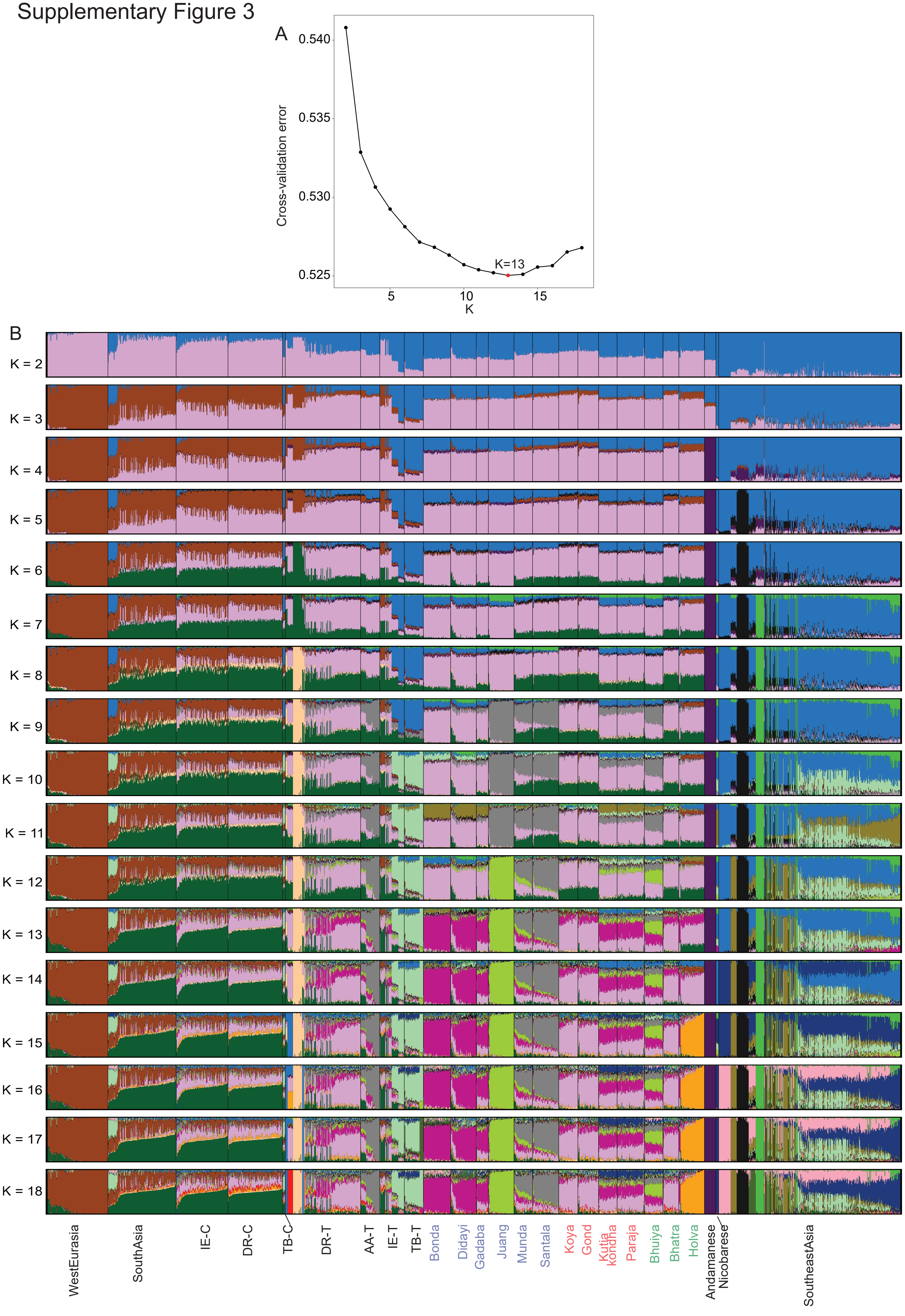

### Supplementary Figure 4

#### Supplementary Figure 4

A

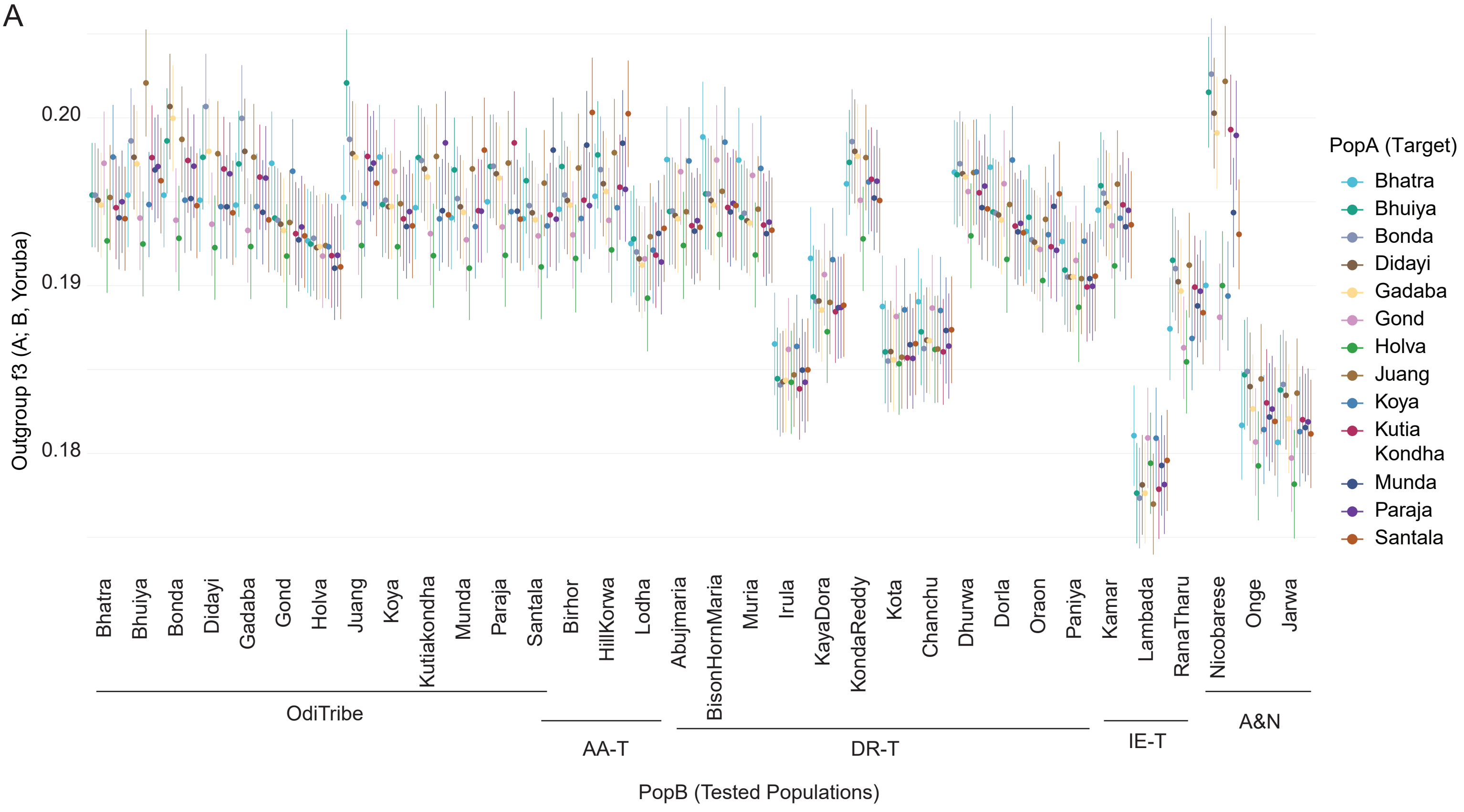


B

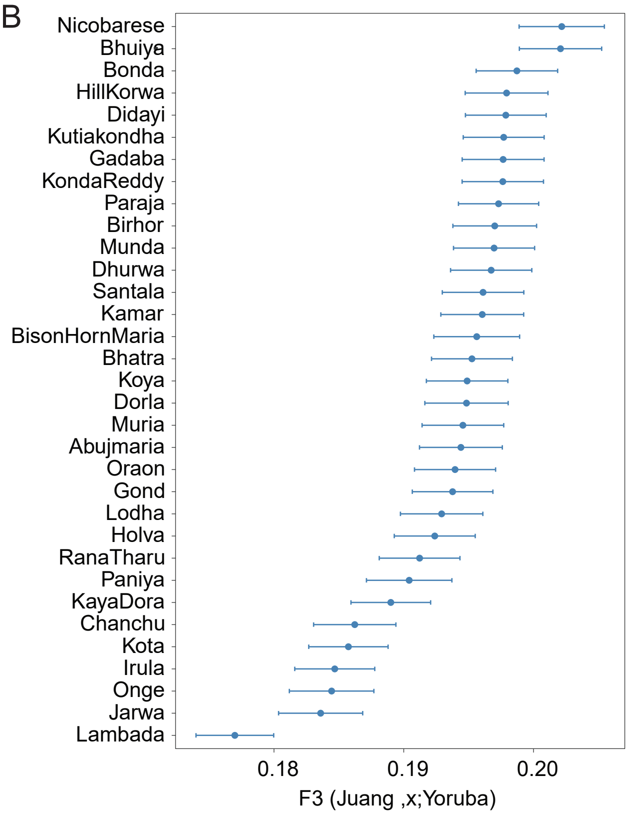


C

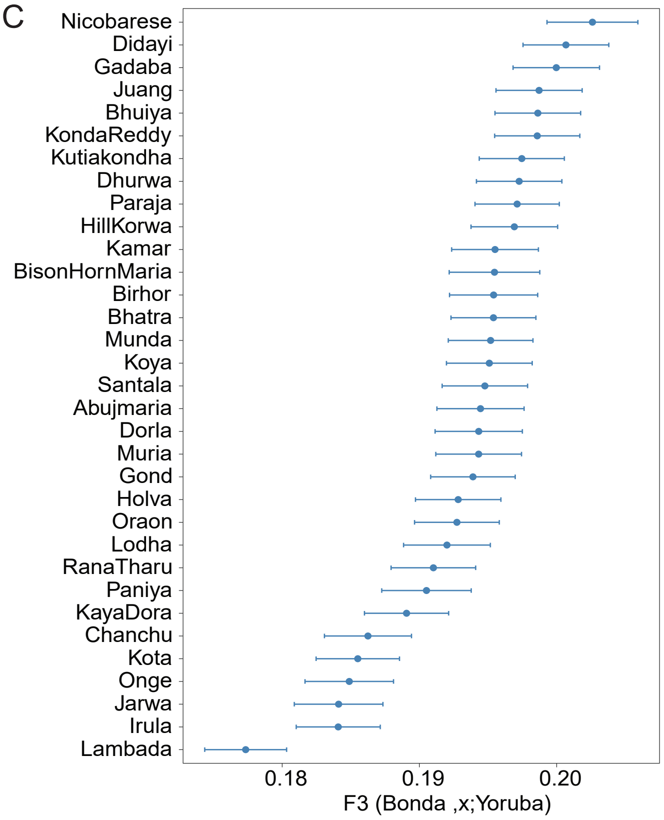

### Supplementary Figure 6

#### Supplementary Figure 6

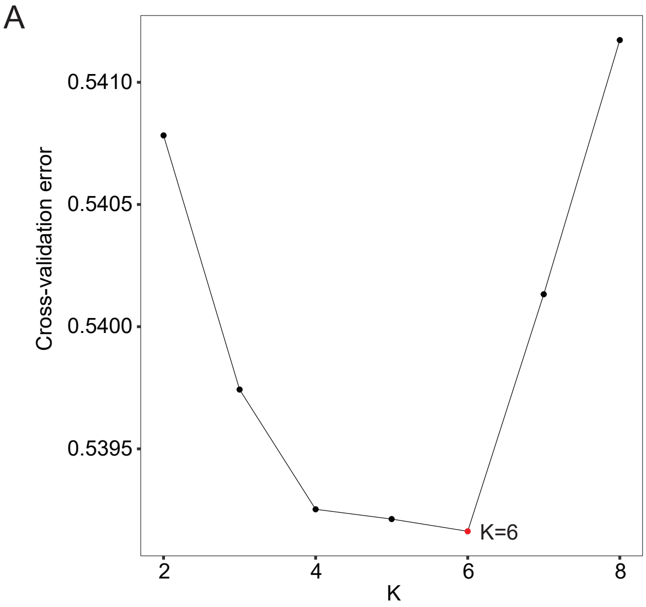


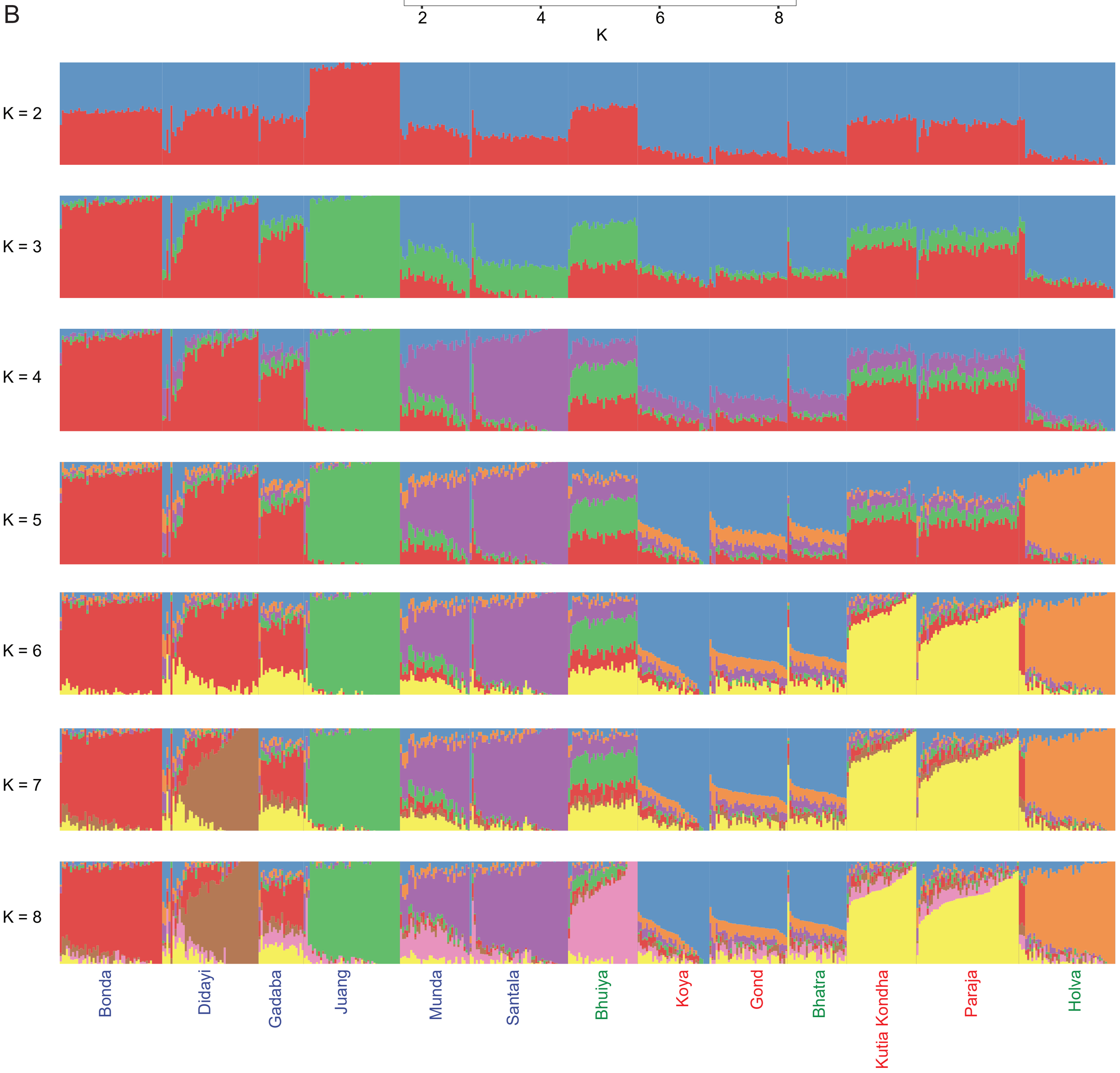

### Supplementary Figure 7

Supplementary Figure 7

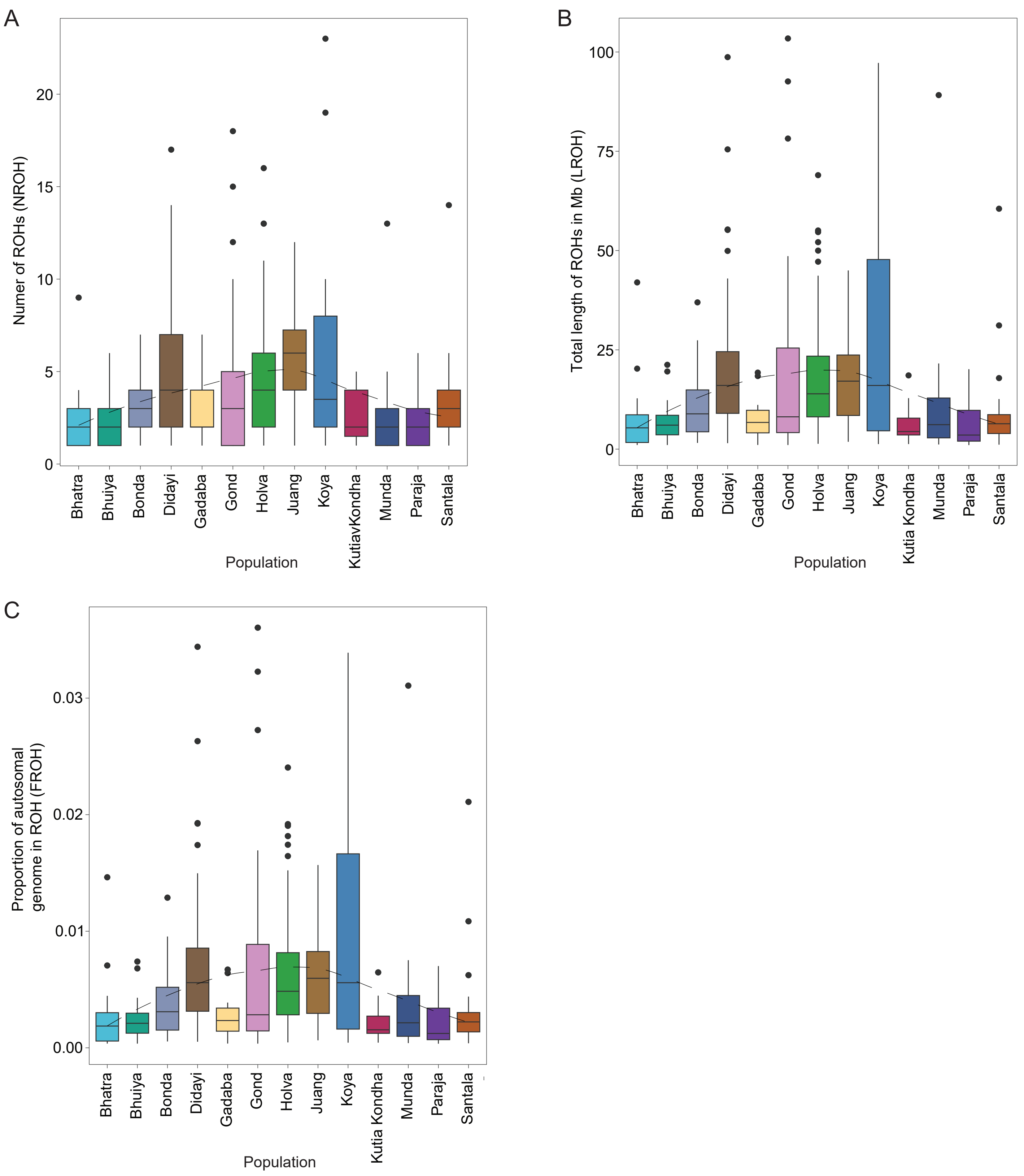

### Supplementary Figure 8

#### Supplementary Figure 8

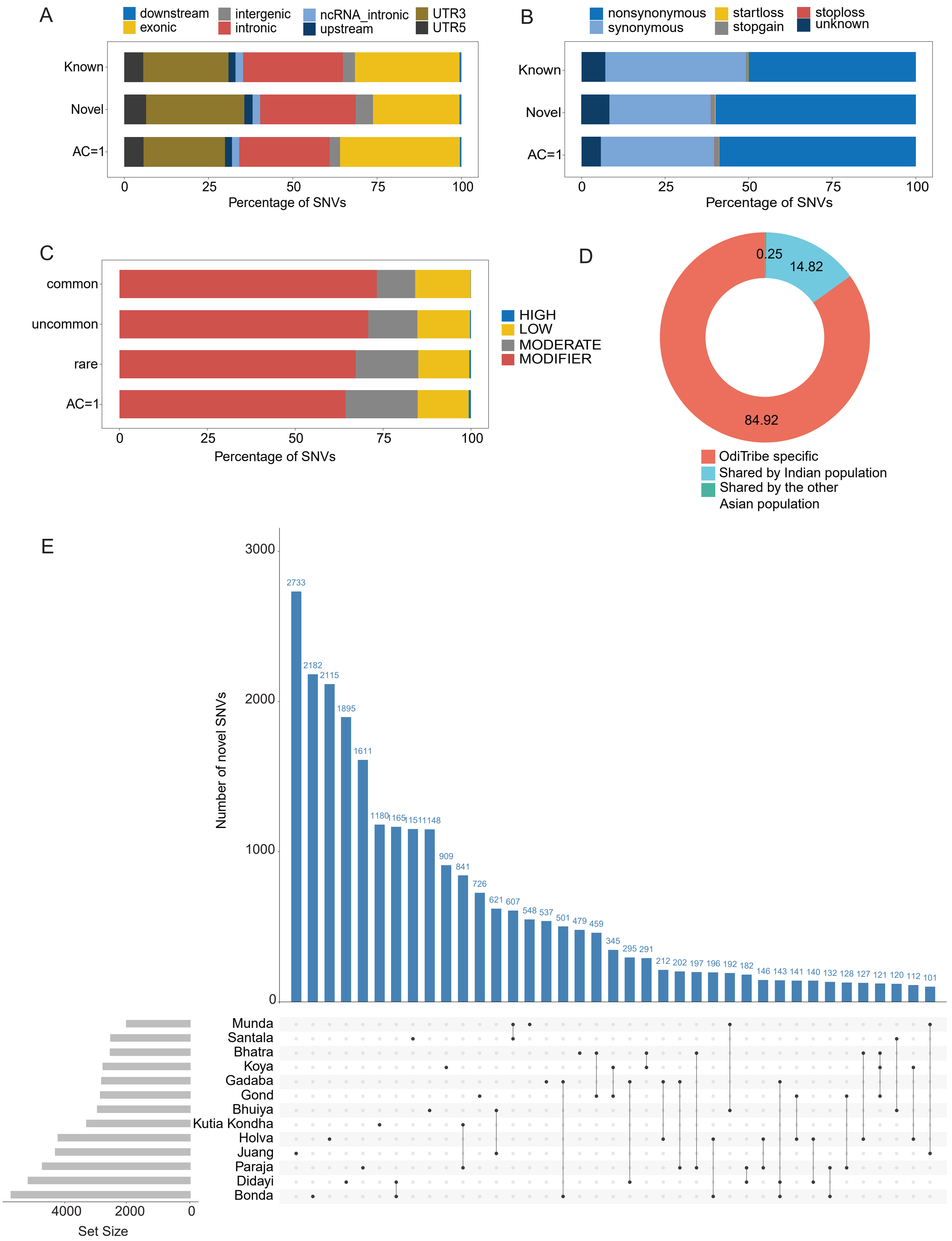

### Supplementary Figure 9

##### Supplementary Figure 9

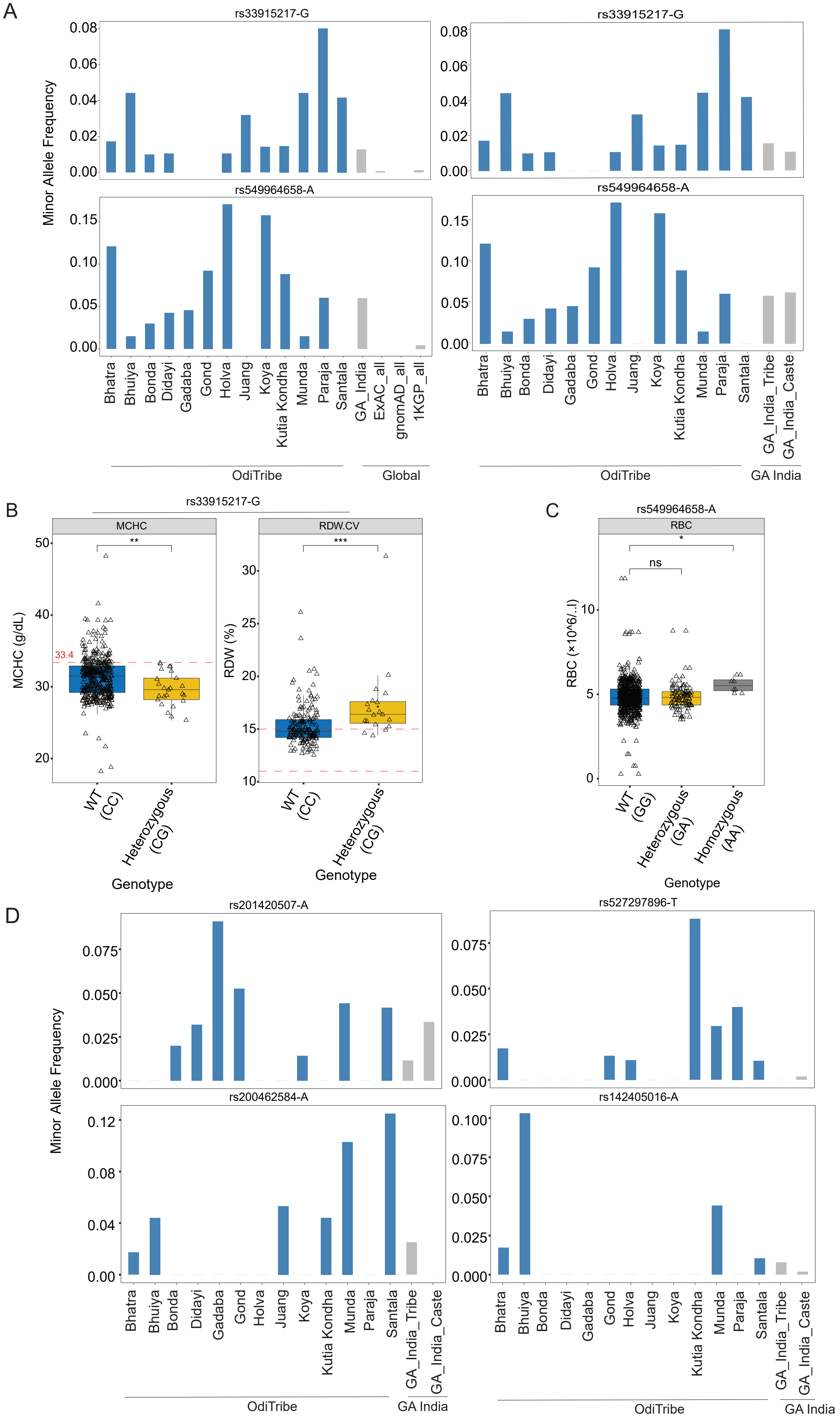

### Supplementary Figure 10

#### Supplementary Figure 10

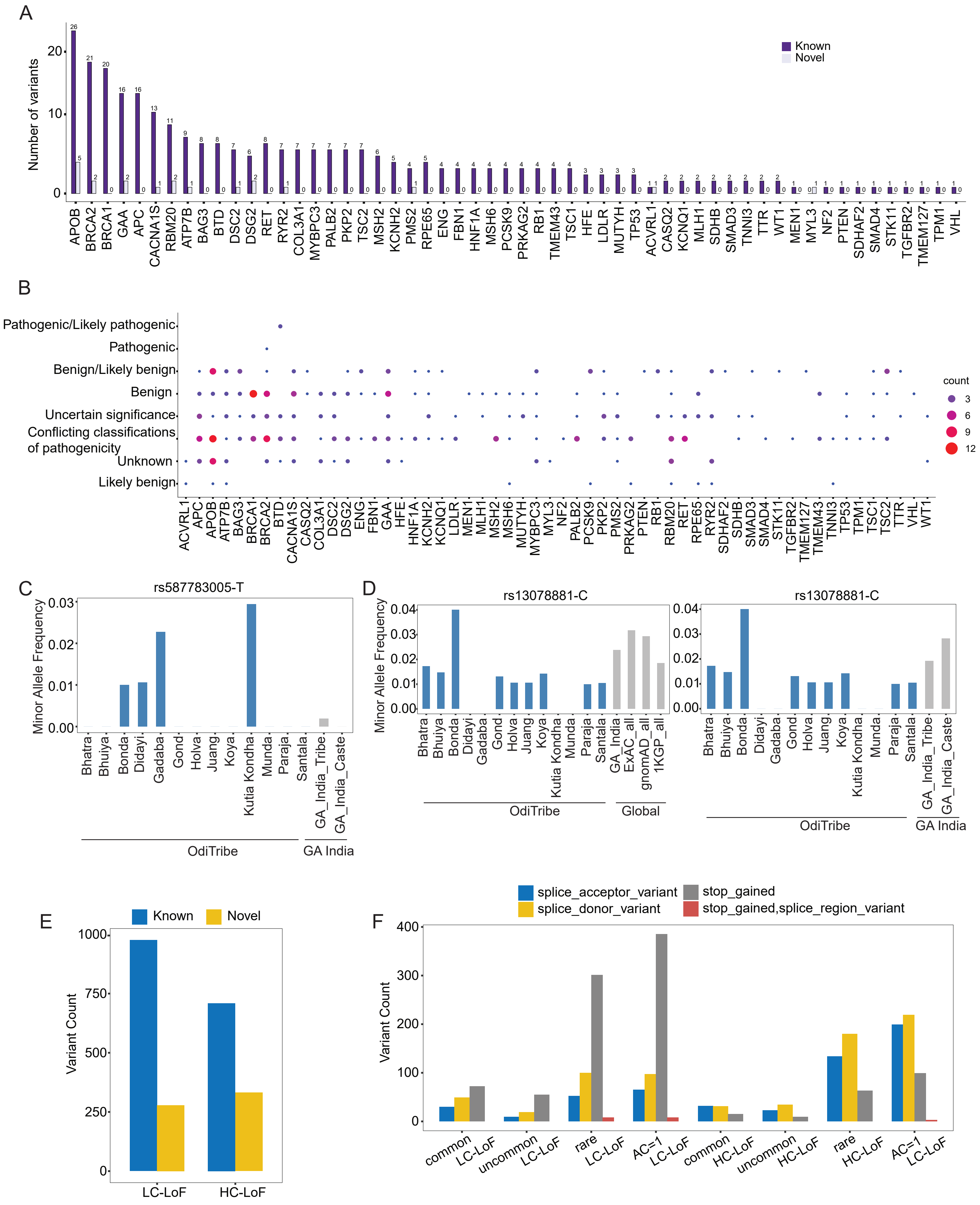
