## Supplementary Figure 5 for "Fine-scale genetic structure and population-specific clinically relevant variants in the indigenous tribal populations of India"

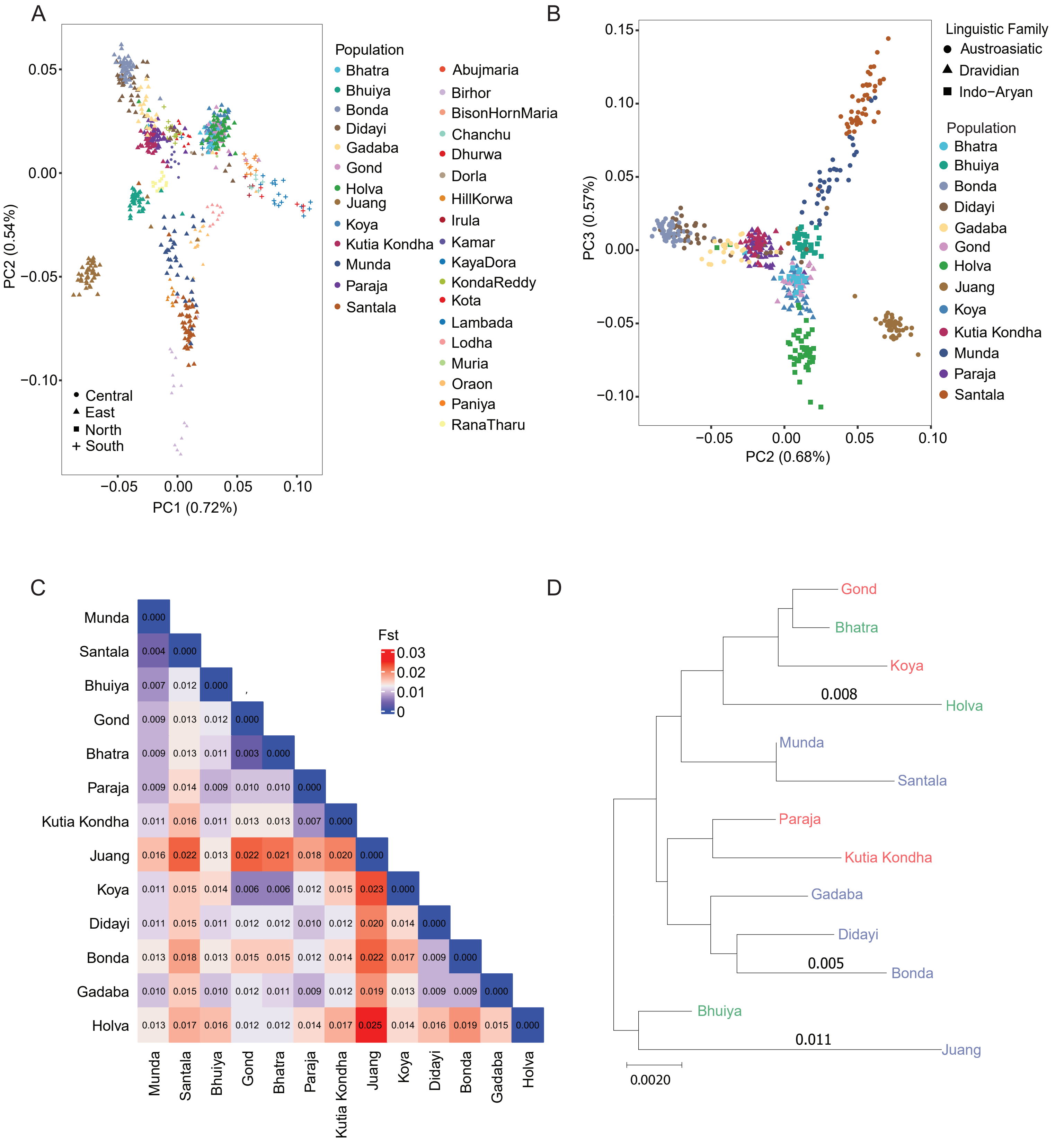


| | Munda | Santala | Bhuiya | Gond | Bhatra | Paraja | Kutia Kondha | Juang | Koya | Didayi | Bonda | Gadaba | Holva |
|---|---|---|---|---|---|---|---|---|---|---|---|---|---|
| Munda | 0.000 | | | | | | | | | | | | |
| Santala | 0.004 | 0.000 | | | | | | | | | | | |
| Bhuiya | 0.007 | 0.012 | 0.000 | | | | | | | | | | |
| Gond | 0.009 | 0.013 | 0.012 | 0.000 | | | | | | | | | |
| Bhatra | 0.009 | 0.013 | 0.011 | 0.003 | 0.000 | | | | | | | | |
| Paraja | 0.009 | 0.014 | 0.009 | 0.010 | 0.010 | 0.000 | | | | | | | |
| Kutia Kondha | 0.011 | 0.016 | 0.011 | 0.013 | 0.013 | 0.007 | 0.000 | | | | | | |
| Juang | 0.016 | 0.022 | 0.013 | 0.022 | 0.021 | 0.018 | 0.020 | 0.000 | | | | | |
| Koya | 0.011 | 0.015 | 0.014 | 0.006 | 0.006 | 0.012 | 0.015 | 0.023 | 0.000 | | | | |
| Didayi | 0.011 | 0.015 | 0.011 | 0.012 | 0.012 | 0.010 | 0.012 | 0.020 | 0.014 | 0.000 | | | |
| Bonda | 0.013 | 0.018 | 0.013 | 0.015 | 0.015 | 0.012 | 0.014 | 0.022 | 0.017 | 0.009 | 0.000 | | |
| Gadaba | 0.010 | 0.015 | 0.010 | 0.012 | 0.011 | 0.009 | 0.012 | 0.019 | 0.013 | 0.009 | 0.009 | 0.000 | |
| Holva | 0.013 | 0.017 | 0.016 | 0.012 | 0.012 | 0.014 | 0.017 | 0.025 | 0.014 | 0.016 | 0.019 | 0.015 | 0.000 |
